# Convolutional neural networks quantify antibiotic resistance in *Mycobacterium tuberculosis* with diagnostic grade accuracy and predict treatment response

**DOI:** 10.1101/2025.08.05.25333066

**Authors:** Sanjana G. Kulkarni, Anna G. Green, Brendon C. Mann, Samantha Malatesta, Suchitra Kulkarni Goodwin, Nina Cesare, Shandukani Mulaudzi, Noorjahn Rawoot, MIC-ML Consortium, Robin Warren, Karen R. Jacobson, Maha R. Farhat

## Abstract

There is considerable interest in training machine learning (ML) models on genomic data that achieve clinical grade diagnostic accuracy. Many successful ML models have been trained and validated on binary tasks because predicting biomedically relevant continuous variables is difficult to optimize. In this work, we present convolutional neural networks (CNNs) that predict minimum inhibitory concentrations (MICs) for eight antibiotics from *Mycobacterium tuberculosis (Mtb)* gene sequences. By including evolutionary information, protein biochemical properties, and data augmentation for rare variants, we build models that predict 89% of MICs within one drug concentration doubling. Although trained on ≤ 52% of the World Health Organization’s (WHO) drug resistance mutation catalogue data, the CNNs accurately predict the effects of 97% of its graded mutations. In a cohort of 373 patients with rifampicin-susceptible *Mtb* infections, higher CNN-predicted rifampicin MICs are associated with unfavorable treatment outcomes, suggesting that subtle differences in MIC below the resistance threshold are clinically relevant. These results demonstrate the value of encoding multiple dimensions of biological data in machine learning of function or cellular phenotypes and that domain knowledge-inspired machine learning models can be both interpretable and reach clinical grade accuracy.

## Introduction

Pathogen genetic data promises to accelerate the diagnosis of antibiotic resistance in infectious diseases.^1,2^ The prediction of antibiotic resistance from pathogen sequence data is well motivated biologically as its almost entirely genetically encoded.^3,4^ For *Mycobacterium tuberculosis* (*Mtb*), point-of-care pathogen genetic assays currently aid clinicians in identifying resistance to rifampicin within a few hours of sample collection.^5,6^ However gaps currently remain in the prediction of resistance to other drugs, including novel drugs like bedaquiline. These may be addressed by synthesizing genetic variant data from whole genome sequencing (WGS) using statistical models^7–11^. Specifically, machine learning (ML) models achieve the highest accuracy by considering non-linear, pairwise and higher order relationships.^12^

Convolutional neural networks (CNNs) are a subset of ML models well suited to prediction from genetic sequence inputs^13–15^. CNNs can be trained directly on the sequence alignment without the need to characterize specific mutations *i.e.* defining individual variants, circumventing challenges with genetic annotation such as encoding multiple frameshift mutations on the same gene.^16^ CNNs can learn the local context around each genetic variant to make accurate predictions on variants that are not present in the training data boosting power and generalizability.

Drug resistance is typically diagnosed as present or absent at an expert-determined drug “critical concentration” (CC).^7,17^ Despite this convention of convenience, antibiotic resistance is a continuous phenotype, binary tests can misclassify bacterial samples that lie close to the CC.^18^ CCs are also periodically revised depending on the changing distribution of clinical resistance globally.^19^ Current data suggests that low levels of resistance, even those below the CC, can increase the likelihood of complete drug resistance developing during treatment^20–22^ and have been found to associate with TB relapse.^23^ However low level resistance is generally only detectable by measuring MICs, which are not performed in clinical practice due to high labor and material cost, biohazard and low precision.^24^ Predicting MICs directly from the genetic sequence rapidly could enable tailored TB therapy, reduce treatment failure, and inform antibiotic stewardship globally.

In this work, we introduce interpretable state-of-the-art CNN models that rapidly and accurately predict MICs from WGS data.^7^ When supplemented with amino-acid translations, biophysical properties, and lineage information, genetic CNNs demonstrate higher accuracy and generalizability. CNNs predict MICs with accuracy on par with *in vitro* growth based drug susceptibility tests and are superior to binary models for predicting treatment outcome in TB. This study offers a large advance in the use of *Mtb* genotypic data to guide TB treatment, as well as methodological insights generalizable to other contexts of ML on sequence data.

## Results

### A. WGS data and MIC distribution

Data from 14,834 *Mtb* isolates with MICs **(Supplementary Data 1)** were obtained from the Comprehensive Resistance Prediction for Tuberculosis: An International Consortium (CRyPTIC) project,^25^ the National Center for Biotechnology Information database, PATRIC,^26^ the Harvard TB CETR,^4^ the 2^nd^ edition of the World Health Organization Resistance Mutation Catalogue,^16^ and unpublished sources curated by members of the MIC-ML consortium **(Supplementary Data 2)**. We focused on MICs for the four first-line anti-tubercular drugs – rifampicin, isoniazid, ethambutol, and pyrazinamide – and four second-line drugs – levofloxacin, moxifloxacin, ethionamide, and bedaquiline. Per drug, sample size ranged from 1,021 to 12,069, and the frequency of resistance (MIC > critical concentration) ranged from 9.6% to 55% **(Table 1, Extended Data Fig. 1)**. MICs measured in different media were scaled to a single medium for each drug.^4^ The lineage distribution is consistent with the global distribution of lineages **(Fig. 1a)**. For pyrazinamide, because of low MIC sample sizes especially in the resistant range, we trained a separate CNN on binary phenotypes **(Extended Data Fig. 3, Methods Section B).** Including this binary data, the sample size ranged from 7,800 to 14,107 across the 8 drugs **(Table 1).**

**Figure 1.**
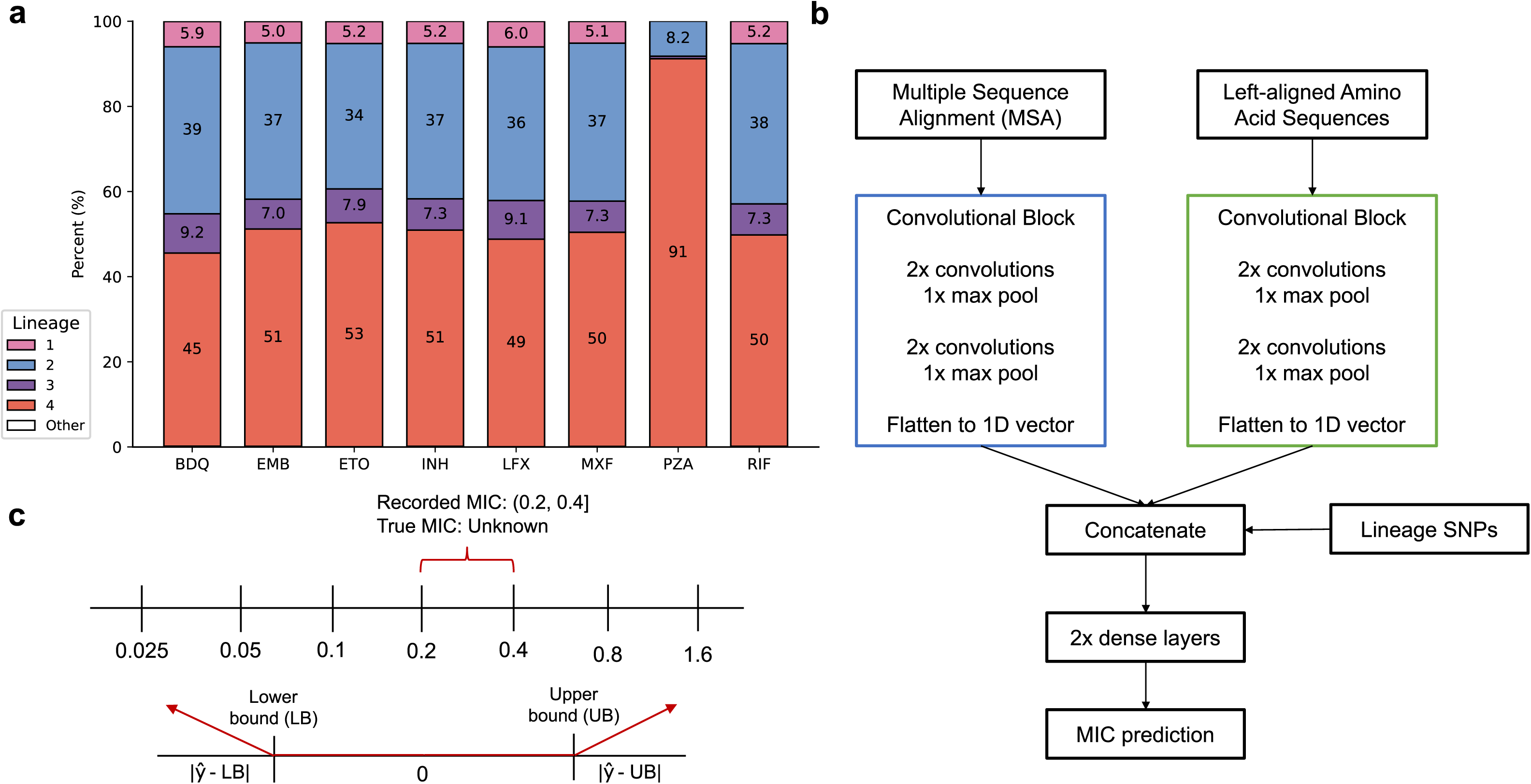
Data and CNN overview. **a:** Multi-modal CNN with two convolutional blocks and inclusion of lineage-defining SNPs. **b**: Custom loss function, which only penalizes predictions outside of the associated MIC range for each prediction, using mean absolute error. **c:** Lineage breakdown of the input datasets for the four most common lineages in the data. The “Other” category is composed of *M. bovis* and L6 isolates, which together are present in fewer than 1% of isolates in each dataset. Pyrazinamide MICs were not measured by the CRyPTIC study,^25^ so the distribution of lineage is highly skewed and not representative of the worldwide lineage distribution.^28^

**Table 1.**
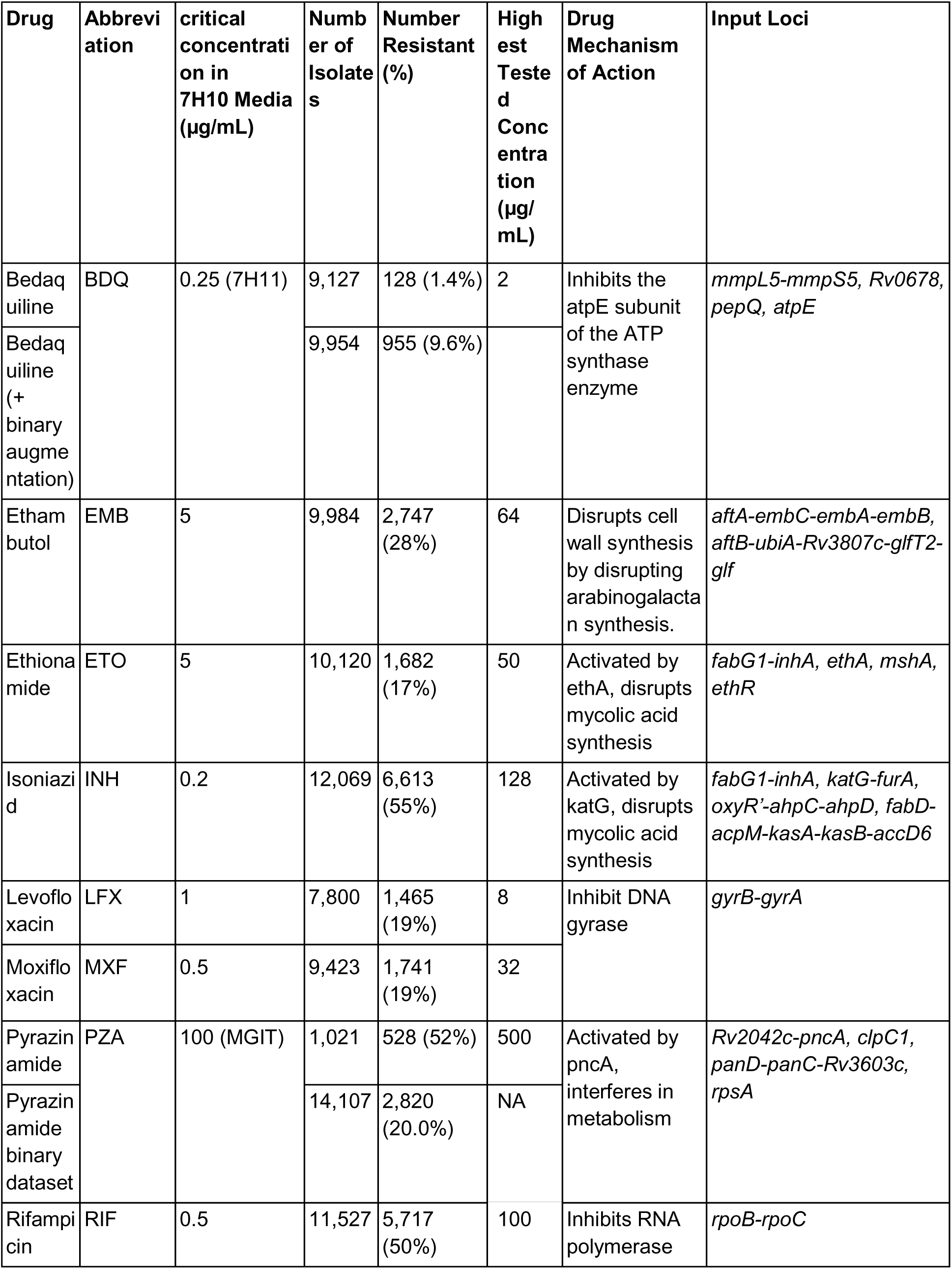
Phenotypic summaries and genetic loci used to train models for each of 8 anti-TB drugs. MIC ranges and averages are computed using the midpoints of the MIC ranges of the isolates. MICs for pyrazinamide were in MGIT medium because all isolates were tested MGIT, MICs for bedaquiline were normalized to 7H11 medium, and all other drug MICs were normalized to 7H10 medium. Due to right-censoring, the largest MICs are greater than the highest tested concentration, but are recorded as, for example, “>2 µg/mL”. We included upstream promoter regions up to the nearest predicted transcriptional start site ^77^ for all loci. Contiguous genes in a single locus are delimited by hyphens, and different loci are delimited by commas. *ethA* and *ethR* were encoded separately due to differing strand senses, but the overlapping intergenic region was included in both loci in each pair. Coordinates and gene names were extracted from Mycobrowser **(Supplementary Table 1)**.^78^ The full list of isolates and their MICs is in **Supplementary Data 1.**

### B. CNN architecture with multimodal input improves prediction accuracy

We trained single-drug CNN models, and for optimal interpretability and computational tractability, relevant core genes were selected as input **(Supplementary Table 1)**.^16^ We implemented a custom error function to accommodate the semi-continuous nature of MICs, including right and left-censoring at the highest and lowest tested drug concentrations **(Fig. 1c, Methods Section G).** Initially we trained CNNs only on the nucleotide sequence, however this training resulted in models that did not sufficiently distinguish nonsense from missense mutation effects (e.g. for *pncA*, **Supplementary Results Section A)**. Inspired by EVEscape,^27^ we added molecular weight (g/mol), isoelectric point, and hydrophobicity (Eisenberg scale) for each amino acid in the proteins encoded by the relevant genes and passed them through independent convolutions **(Fig. 1b).** This multimodal input allowed the CNNs to learn from both the nucleotide and protein sequence, their amino acid characteristics, and implicitly, the translated protein length. The multimodal models were able to distinguish between the effects of nonsense and missense mutations **(Extended Data Fig. 3)**.

### C. Incorporation of lineage boosts performance

*Mtb* is a clonal bacterium that separates into 10 major genetically distinct populations or lineages^28^. Lineage defines hundreds to thousands of linked variants across the genome that can encode MIC effects or interact with canonical resistance mutations.^29^ We tested if adding lineage information using a validated 62 silent mutation barcode^30^ improves resistance prediction accuracy **(Methods Sections F-G)**.

Lineage information significantly improves model performance for bedaquiline, ethambutol, ethionamide, levofloxacin, and moxifloxacin (one-sided Welch’s t-test p < 0.04, mean absolute error (MAE) decrease of 0.004-0.09 log2-MIC units in cross-validation), but significantly decreased pyrazinamide MIC performance (p = 0.02, MAE increase of 0.07 in cross-validation) due to the small pyrazinamide dataset size and strong lineage skew **(Fig. 2a-h)**. For the binary pyrazinamide CNN, the lineage barcode did not significantly affect performance (two-sided Welch’s t-test p = 0.93), possibly due to larger sample size and better lineage representation **(Extended Data Fig. 3)**. The lineage barcode did not significantly alter performance for rifampicin and isoniazid (p ≥ 0.23). The final CNNs include lineage information for all drugs except pyrazinamide MIC. The learned relationships between lineages and MICs are supported by the average MICs by lineage **(Supplementary Results Section D).**

**Figure 2.**
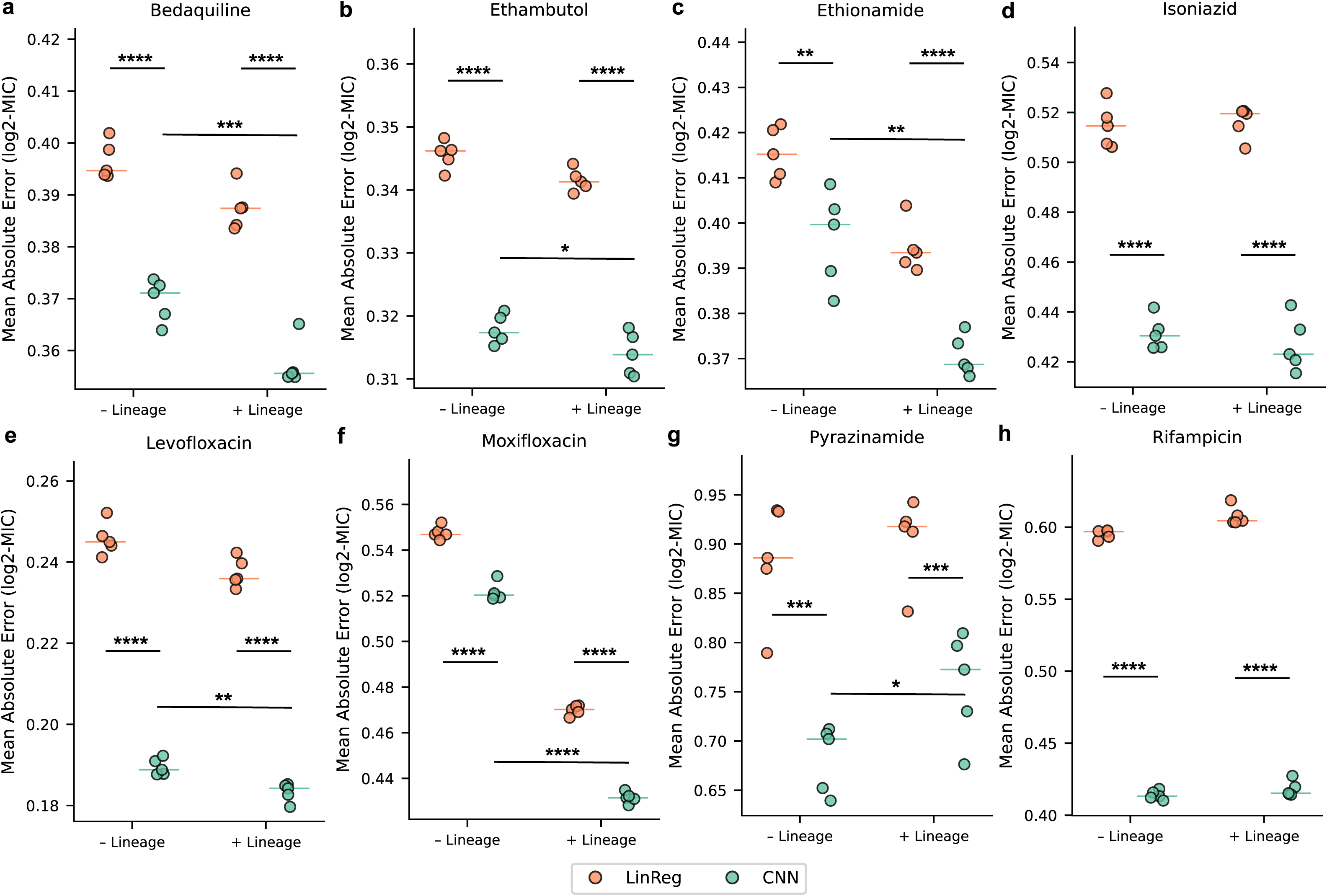
Model metrics between CNN and linear regression on the test set. **a-g**: Mean absolute error of regression models and CNNs for all 7 drugs, with and without lineage SNPs. The units of mean absolute error are log2-MIC units. Replicates are 5 cross-validation splits. y-axes are different across drugs to better show the variability for single-drug models. P-values are one-sided Welch’s t-tests. For readability, p-value bars are only shown comparing performance between CNN models. *: p < 0.05, **: p < 0.01, ***: p < 0.001, ****: p < 0.0001. No annotation indicates not significant.

### D. CNNs outperform regression and predict MIC within 1 doubling for 89% of isolates

To evaluate the value of the CNN architecture, we benchmarked its error against a simpler linear regression approach **(Methods Section G)**. For all 8 drugs, the CNNs achieve significantly lower MAE than regression (one-sided Welch’s t-test p-values < 2 x 10^-4^) **(Fig. 2)**. CNNs’ MAE on the hold-out test sets range from 0.18-0.67 log_2_(MIC) (*i.e.,* MIC doublings). Essential agreement (EA) – the proportion of predictions within 1 doubling dilution of the true MIC – is frequently used to evaluate antimicrobial susceptibility testing and its reproducibility as one doubling is the expected error margin for most drugs.^31^ EA ranged from 79-93% (average 89%) on the test sets for single-drug CNNs, with the lowest being for pyrazinamide **(Supplementary Data 3).** The binary pyrazinamide CNN performed significantly better than logistic regression **(Extended Data Fig. 3)**, with an average AUC difference of 2.5 percentage points (one-sided Welch’s t-test p = 2.4 x 10^-4^).

### E. CNNs achieve state-of-the-art performance

We studied how the CNNs compare with the 2^nd^ version of the WHO *Mtb* resistance mutation catalog,^16^ which was trained on larger datasets ranging from 14,000-48,000 isolates per drug. On the hold-out test sets, we predicted MICs, then dichotomized them at the critical concentrations for each drug to simulate binary resistance prediction. We then compared the sensitivities and specificities to the catalog classification method **(Supplementary Table 2, Fig. 3a-b).** For pyrazinamide, we performed the comparison to the binary CNN because the quantitative model had insufficient training data. Across the drugs, the CNNs have both higher sensitivity and specificity than the catalog (+2.2% and +1.6% on average for sensitivity and specificity, respectively). CNN sensitivities were higher for ethambutol, isoniazid and levofloxacin (p = 2 x 10^-7^, 1x 10^-14^ and 0.01 two proportion z-test), and CNN specificity higher for ethambutol (p = 5.9 x 10^-8^). The CNN sensitivity is lower for ethionamide (p = 8.3 x 10^-4^), with corresponding greater specificity (p = 4.3 x 10^-14^), and otherwise, including for the drug bedaquiline, CNN performances were on par with the catalog classification method (p > 0.05). The essential agreement rates of the CNNs are comparable to those reported by the CRyPTIC using XGBoost models trained on whole-genome k-mers. However the latter models are less interpretable and used non-causal variants in their prediction. They also have substantially lower specificity for bedaquiline resistance prediction **(Supplementary Results Section C)**.

**Figure 3.**
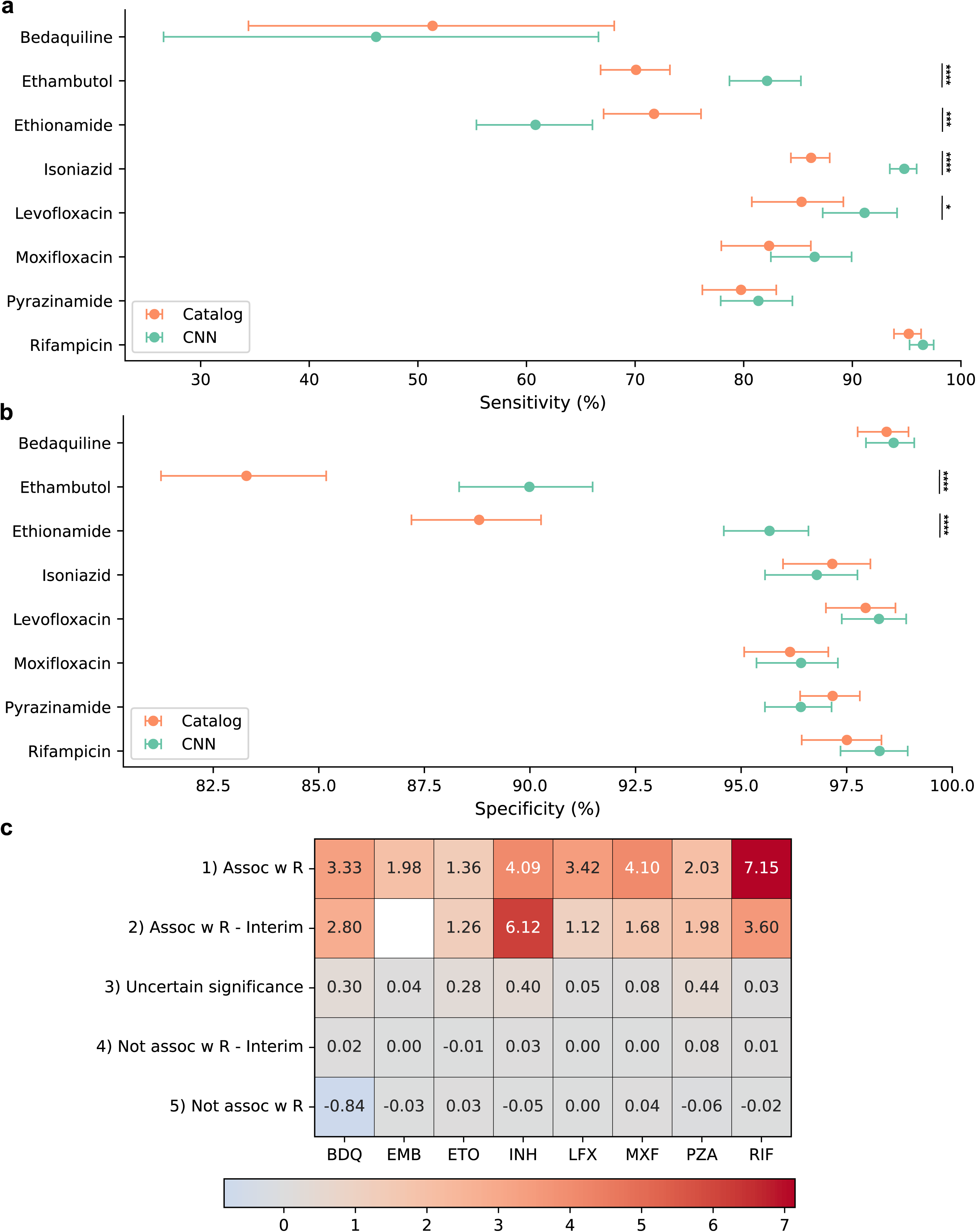
Comparison of binary prediction metrics between the quantitative CNNs and catalog-based classification using the WHO catalog. **a:** Heatmap of the average log_2_(fold changes) of predicted MICs for mutations across the five catalog gradings. **b:** Sensitivity and **c**: specificity of binary resistance predictions of the WHO catalog classification method and the MIC predictions of the CNN, dichotomized at the critical concentrations. 95% confidence intervals (CIs) are exact binomial confidence intervals computed using the Clopper-Pearson method. CI significance testing was performed using the two-proportion z-test. *: p < 0.05, **: p < 0.01, ***: p < 0.001, ****: p < 0.0001. No annotation indicates not significant. Source data are in **Supplementary Data 6.**

### F. CNNs attribute resistance to specific variants accurately and exhaustively

To validate the variants the CNNs learned from, we created synthetic sequences with all substitutions and in-frame indels in the WHO TB drug resistance mutation catalog^16^ and predicted MICs for them using the pre-trained models in **Results Section C**. The catalog consists of two categories of resistance-associated mutations and two categories of mutations that are not associated with resistance that differ by the strength of evidence (associated or associated-interim), as well as a fifth uncertain grading for mutations with insufficient evidence to be classified. The average predicted MIC (transformed to log_2_FC) for mutations in each catalog grading correlated with the strength evidence and direction of association for that grade **(Fig. 3c, Supplementary Data 4)**, including a higher average MIC for “associated with resistance” than for “associated with resistance interim.” The exception was isoniazid, for which the category with the largest predicted average MIC was the ‘interim’ category. This was because this category contains many *katG* truncations graded interim due to rarity but have higher predicted MICs than *katG* substitutions that predominate in the “associated with resistance” category. For the 28 mutations in the WHO catalog with literature evidence against drug resistance association,^32,33^ the average CNN-predicted log_2_FC was 0.02 (range = -0.73-0.89) **(Supplementary Data 4)**. For the binary pyrazinamide CNN, we computed the proportion of variants in each category that were predicted to associate with resistance. This proportion is highest for Group 1 at 96% and decreases monotonically to Group 5 at 0% **(Supplementary Data 4)**.

### G. CNNs identify mutations associated with subtle shifts in MIC

A key advantage of predicting resistance quantitatively is the identification of borderline or intermediate effects on antibiotic resistance. In the WHO catalog, 25 variants are associated with intermediate resistance – 13 moxifloxacin, 8 isoniazid, and 4 rifampicin **(Extended Data Fig. 5)**. The MIC distribution of isolates with borderline resistance variants often straddle the critical concentration overlap with the susceptible and lower resistance range,^34–36^ and the CNNs reliably predict these intermediate effects **(Extended Data Fig 5)**. For 7 of the 8 isoniazid variants (1 is absent from the training data and not predicted to elevate MIC).^37,38^ García-Marín *et al^39^* identified katG_p.Leu48Pro and katG_p.Gly273Arg as additional putative borderline isoniazid resistance variants (not in the WHO catalog). The CNN predicts MICs of 0.20 and 0.30 µg/mL for katG_p.Leu48Pro and katG_p.Gly273Arg, respectively, in agreement with the *in vitro* results. For moxifloxacin and rifampicin, the 13 and 4 mutations, respectively, are also predicted to have intermediate resistance by the CNN (predicted MICs of 0.10-1.9 µg/mL for moxifloxacin, and 0.33-1.9 µg/mL for rifampicin (>1 only for rpoB_p.Ile491Phe, **Extended Data Fig 5)**.

### H. CNNs learn drug binding pockets and novel antibiotic resistance variants in both essential and non-essential genes

Because of the convolutional component, CNNs can potentially learn the protein region that impacts drug binding or effect. We confirm this and assess the interpretability of the model by studying the 3D distribution of mutations predicted to encode high MIC using the Getis-Ord (GO) clustering statistic.^40^ GO is computed using the predicted log_2_FC in MIC for each residue and their pairwise distance in the protein. We tested for significant clustering of high or low levels of resistance around each residue using a permutation test **(Methods Section J)**.

The CNN predictions demonstrate significant clustering around known drug binding pockets (also the active sites) in the essential drug targets RpoB **(Fig. 4a)** and GyrA and GyrB **(Fig. 4b)**. All 27 residues of the rifampicin resistance determining region (RRDR) (coordinates 426-452) in *rpoB* are clustered (*a.k.a.* “hot spots”), and an additional 62 hot spots occur between residues 48 and 674 but are all located near the RRDR in 3D space **(Fig. 4a).** Similarly, we observe 55 hotspots in the gyrase subunits near the moxifloxacin- and DNA-binding sites, 35 of which are in the quinolone resistance determining regions of *gyrA* and *gyrB* **(Fig. 4b)**.^41^ We do not observe hotspots outside of these active sites for essential proteins.

**Figure 4.**
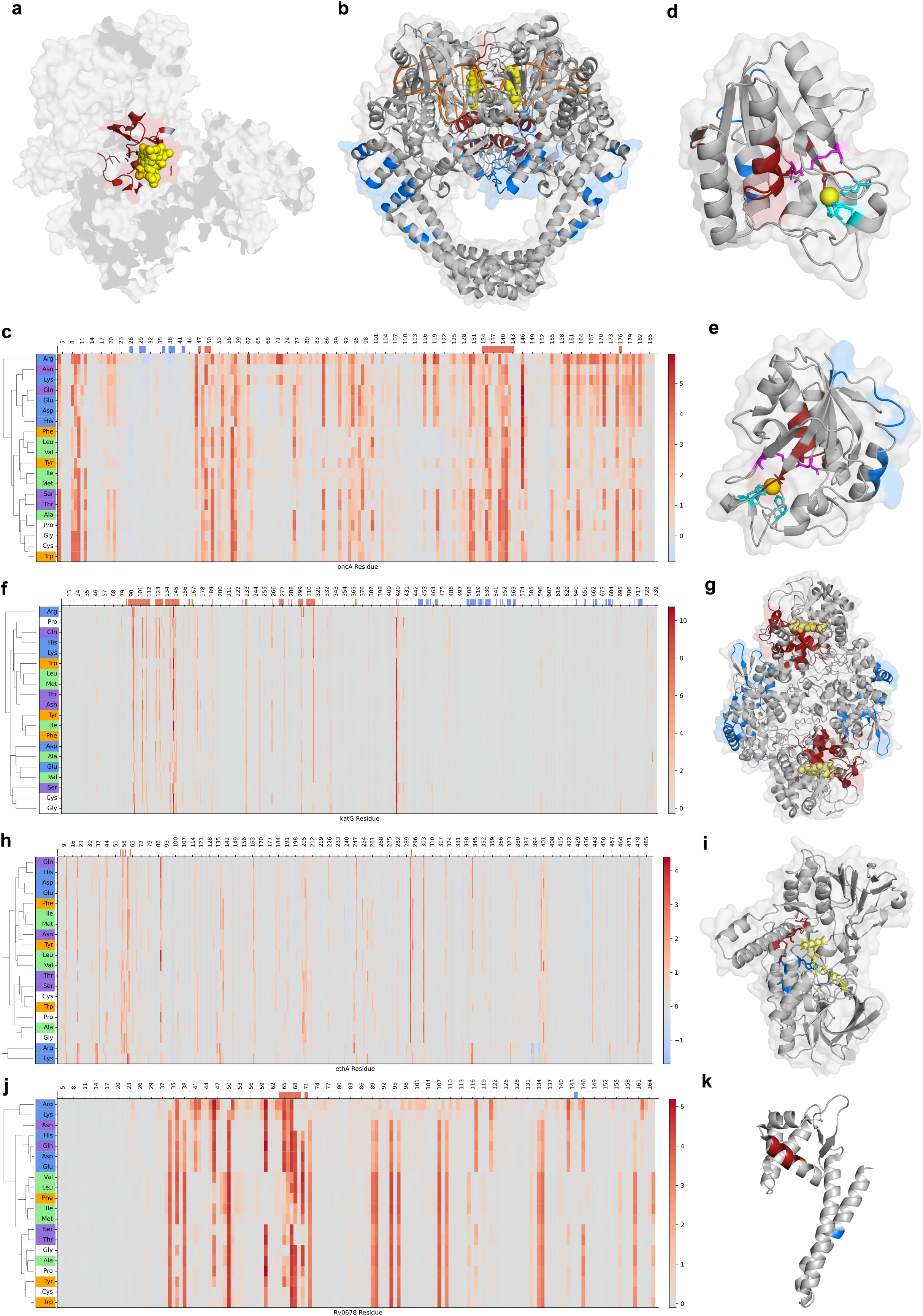
*In silico* predictions for missense mutations identifies regions of both essential and non-essential genes that are highly associated with resistance. For all protein structures, hot spot residues are colored red, and cold spot residues are colored blue. **a:** Hotspots in rpoB. Yellow = rifampicin. There are no cold spots in rpoB. **b:** Hot and cold spots in the gyrA-gyrB complex. Yellow = moxifloxacin, orange = DNA. **c, f, h, j:** Heatmaps of MIC log_2_FC for missense mutations, except residue 1, with hierarchical clustering on the amino acid axis for pncA **(c)**, katG **(f)**, ethA **(h),** and Rv0678 **(j)**. The top x-axis of the heatmaps shows hot and cold spots with significant clustering in 3D space. **d-e:** Two views of pncA with hot and coldspots. Yellow = Fe^2+^ coordinating ion. Cyan = Iron-coordinating residues (49, 51, 57, 71), one of which is a hotspot (red). Pink = catalytic triad, one of which is a hotspot (red). The side chains of the 4 iron-coordinating residues and the catalytic triad residues are shown. **g:** katG homodimer with heme substrates (yellow). **i:** ethA AlphaFold predicted structure. Yellow = predicted FAD binding site by AlphaFill. Red = Hot spots from GeO clustering, blue = residues 391, 393, and 397, which have the lowest average predicted MIC fold changes among missense mutations. **k:** Hot-(red) and coldspots (blue) in the Rv0678 transcription factor. The hot spots occur in the DNA-binding domain, and the single cold spot in the dimerization domain. Only a single monomer is shown for readability.

We observe similar structural clustering in non-essential proteins that are not drug targets. These include PncA and KatG, which activate the pro-drugs pyrazinamide and isoniazid, respectively, and Rv0678, a transcription factor that represses expression of bedaquiline efflux pumps mmpL5 and mmpS5 (ref). In PncA, the clustering occurs in the pyrazinamide- and coordinating iron-binding sites **(Fig. 4d-e)**. In KatG, the clustering separates between the termini. The hot spot associated with elevated MIC occurs near the heme binding site that mediates drug activation in the N-terminus. The hot spot in the C-terminus is associated with lower MIC. The C-terminus is likely important for protein stability and homodimer formation because C-terminus deletions also lead to INH resistance.^35^ **(Extended Data Fig. 3f).** In Rv0678, hot spots occur at residues 64-71 in a single helix of the DNA-binding domain of each monomeric subunit **(Fig. 4k)**.^42^ The full DNA-binding domain extends from residues 34-99, and many substitutions within are predicted to elevate MICs **(Fig. 4j)**. A single cold spot occurs in 3 of the 4 monomers in the dimerization domain. Mutations in the Rv0678 hot spot likely inhibit or reduce DNA binding upstream of the bedaquiline efflux pumps, resulting in higher efflux pump transcription and bedaquiline resistance.

We hypothesized that CNN prediction clustering can characterize new structure-function relationships for proteins known to mediate resistance without a well characterized mechanism. The non-essential gene *ethA*, encodes an FAD-dependent mono-oxygenase that activates the prodrug ethionamide. Disruptive mutations in *ethA* are the major source of resistance-causing mutations.^35^ However, without a solved protein structure, the exact binding sites of EthA and its mechanism of drug activation are not known. We overlaid CNN predictions on the AlphaFold confident substructure of EthA (residues 2-483 of 489 total residues with high structure confidence scores, pLDDT > 70). In parallel, we predicted the binding pocket of FAD using AlphaFill, which relies on homology to ligand- and cofactor-binding sites in protein complexes.^43^ The predictions placed FAD within 3 Å of the nearest high MIC cluster in ethA **(Fig. 4i)**. These results highlight the ability of machine learning models to identify structurally relevant regions of proteins without being explicitly trained on structural information.

### I. CNNs identify novel resistance associations with ethionamide and other drugs

The WHO mutation catalog grades any loss of function (LoF) mutation in *ethA* as resistance associated by expert opinion that ethionamide activation from its prodrug to drug form is not possible if the main known activating enzyme EthA is not functional.^16^ However, a substantial number of ethionamide-susceptible isolates with *ethA* LoF mutations have been previously reported,^44,45^ We observe this in in our data **(Extended Data Fig. 6a)** and find a low specificity of the WHO catalog for ethionamide resistance due to the *ethA* LoF mutations **(Results Section C)**. Novel putative activators, such as *Rv3083* (*mymA*)^46^, *Rv0565c^45^*, *mshA^47–49^*, *ndh^50^* may directly or indirectly activate ethionamide in isolates with *ethA* LoF allowing for continued susceptibility to the drug. To study the role of these genes, we built an expanded model with the WHO Tier 2^16^ genes *Rv0565c, Rv3083,* and *ndh* added to the previous Tier 1 loci (*ethA, ethR, fabG1-inhA* and *mshA*). The expanded model had higher albeit not statistically significant accuracy (p=0.11, one sided Welch test), but allowed the identification of 20 novel ethionamide resistance associations in *mshA, Rv0565c, Rv3083,* and *ndh*, all of which are graded uncertain in the catalog **(Table 2, Extended Data Fig 6e-d).** Most (N = 16) are nonsense/LoF mutations **(Extended Data Fig. 6c).** Isolates with a LoF mutation in either *ethA, mshA, Rv3083, Rv0565c,* or *ndh* have significantly higher MIC than isolates with no LoF mutations in each gene **(Extended Data Fig. 6).** The larger the proportion of activator enzymes that harbored an LoF, the higher the ethionamide MIC (3 of 5 with LoFs *vs.* 2 of 5 with LoFs, one-sided Welch’s t-test p = 3.6 x 10^-7^; 2 of 5 vs. 1 of 5 LoFs. p = 3.0 x 10^-3^). LoFs were observed in up to 3 of the 5 ethionamide activator enzymes but were not correlated between the activators (Spearman ρ < 0.40). These results support that multiple LoF mutations in ethionamide activation pathways are required to affect ethionamide resistance.

**Table 2.** 20 mutations in *Rv0565c, Rv3083, mshA,* and *ndh* from the 2023 WHO catalog with a predicted ethionamide MIC greater than 1 doubling relative to the wild-type prediction. All are graded as Group 3) Uncertain confidence in the catalog. Variants are ordered by gene, then by decreasing predicted MIC. The predicted MIC for H37Rv is 1.1 µg/mL, and the critical concentration for ethionamide is 5 µg/mL.

| Gene | Variant | Effect | Predicted MIC (µg/mL) |
| --- | --- | --- | --- |
|  | Rv0565c_p.Tyr146dup | inframe_insertion | 5.8 |
|  | Rv0565c_p.Trp395* | stop_gained | 3.0 |
|  | Rv0565c_p.Gly189Arg | missense_variant | 2.9 |
|  | Rv0565c_p.Arg462* | stop_gained | 2.8 |
|  | Rv0565c_p.Gly83del | inframe_deletion | 2.7 |
|  | Rv0565c_p.Lys408* | stop_gained | 2.6 |
|  | Rv0565c_p.Gln351* | stop_gained | 2.5 |
|  | Rv0565c_p.Trp266* | stop_gained | 2.5 |
| *Rv0565c* | Rv0565c_p.TrpLeu266* | stop_gained | 2.5 |
| Rv3083 | Rv3083_p.Trp293* | stop_gained | 11.4 |
|  | Rv3083_p.Trp261* | stop_gained | 2.3 |
|  | Rv3083_p.Arg158_His165del | inframe_deletion | 2.3 |
| mshA | mshA_p.Gln128* | stop_gained | 7.7 |
|  | mshA_p.Tyr123* | stop_gained | 7.1 |
|  | mshA_p.Gly311* | stop_gained | 4.8 |
|  | mshA_p.Gln205* | stop_gained | 4.4 |
|  | mshA_p.Gln363* | stop_gained | 3.5 |
|  | mshA_p.Glu220* | stop_gained | 2.9 |
|  | mshA_p.Glu353* | stop_gained | 2.4 |
| ndh | ndh_p.Tyr342* | stop_gained | 3.7 |

Expanding to all 8 drugs, the saliency analysis identifies 247 nucleotides (in 244 unique sites) with significant association with resistance, elevated predicted MIC, are observed at a frequency of at least 5 isolates, and are not currently associated in the WHO catalog **(Supplementary Data 5).** 74% of these are associated with ethionamide and isoniazid resistance, occurring in *ethR* and *katG* respectively, but we also find 5 in *mmpL5* and *Rv0678* related to bedaquiline. The models also identify 23 non-silent mutations from the WHO catalog as associated with hypersusceptibility **(Fig 4h-i**, **Table 3, Supplementary Results Section F).**

**Table 3:**
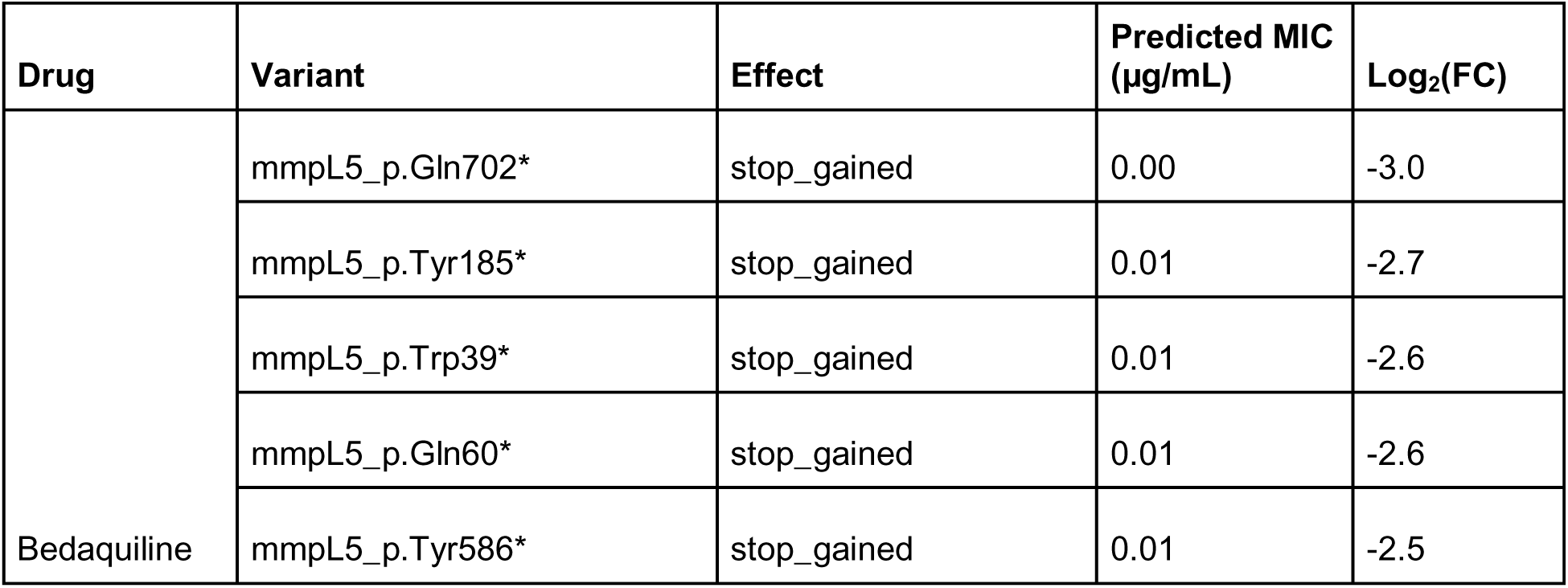
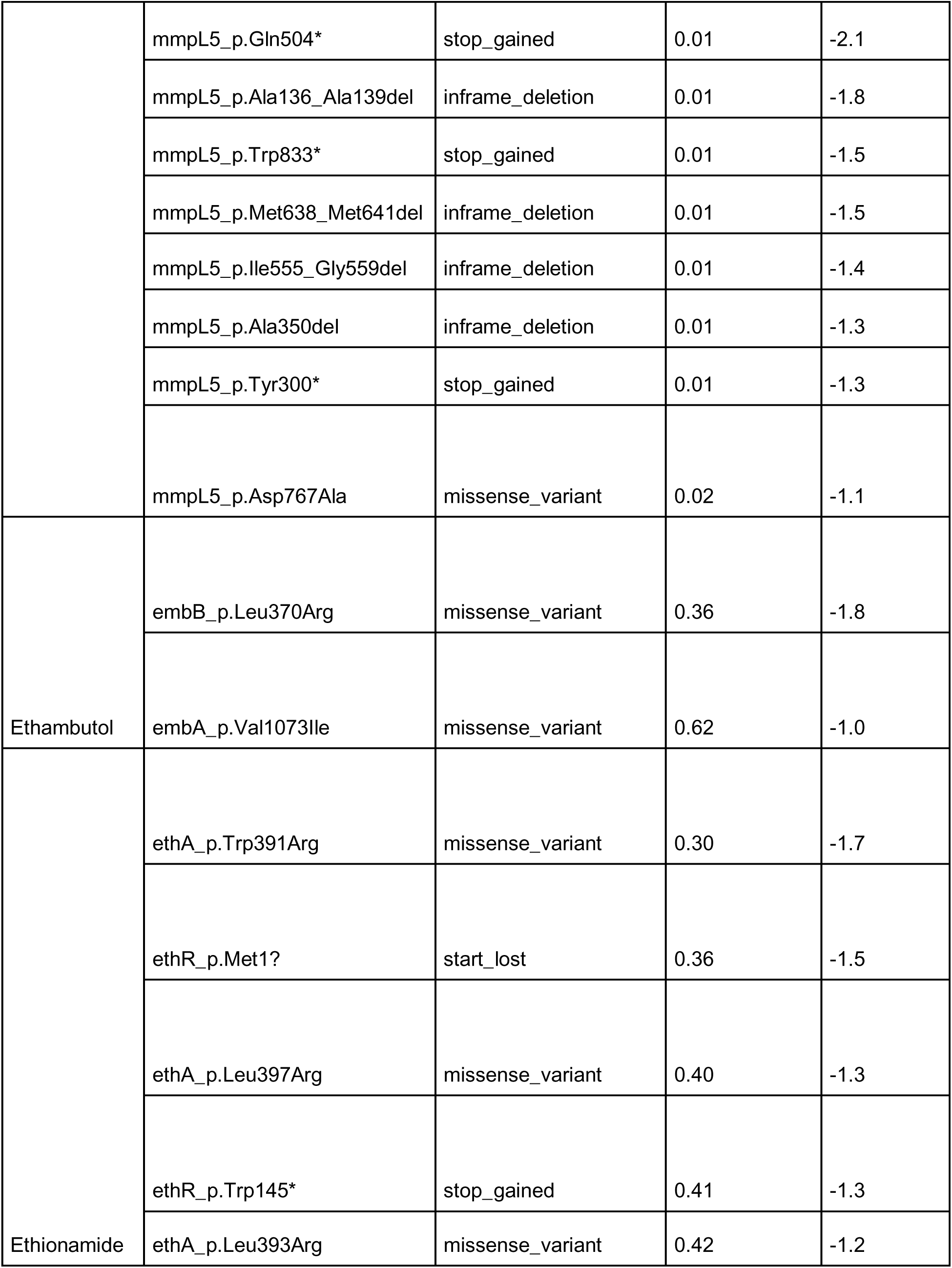
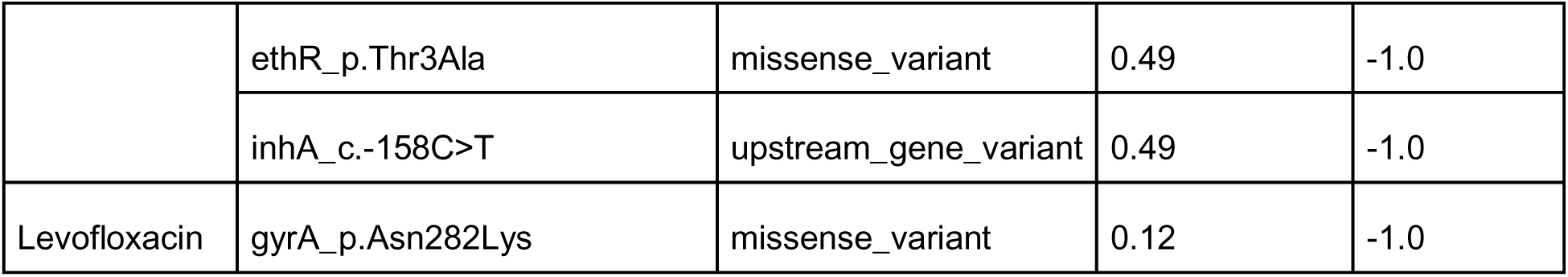
Mutations with evidence for association with drug hypersusceptibility. 23 mutations with predicted log_2_FC ≤ 0.5. Mutations are ordered by drug then by increasing predicted MIC. The predicted MIC for H37Rv (“wild-type”) is 0.03 µg/mL.

### J. CNNs can predict MICs below the critical concentrations with moderate accuracy

Predicting high-level resistance, *i.e.* MICs above the critical concentration, has established value for clinical management of TB. We explore here the accuracy and value of predicting MICs below the critical concentrations in light of a recent study identifying a strong association between sub-critical concentration rifampicin and isoniazid MICs and TB relapse.^23^ We leveraged data from a cohort study of 373 participants with rifampicin-susceptible TB confirmed by the commercial assay GeneXpert-TB/RIF or -Ultra and finely measured MICs **(Methods Section K).**^51^ Of the bacterial isolates collected from 372 participants (one did not have MICs measured, and only 262 had measured pyrazinamide MICs), 25 (7%) had isoniazid resistance, 2 (0.5%) had ethambutol resistance, and 2 (0.8%) had pyrazinamide resistance **(Fig. 5a)**. The MIC measurements span 4 doublings below the critical concentration for each drug **(Extended Data Fig. 7d).**

**Figure 5:**
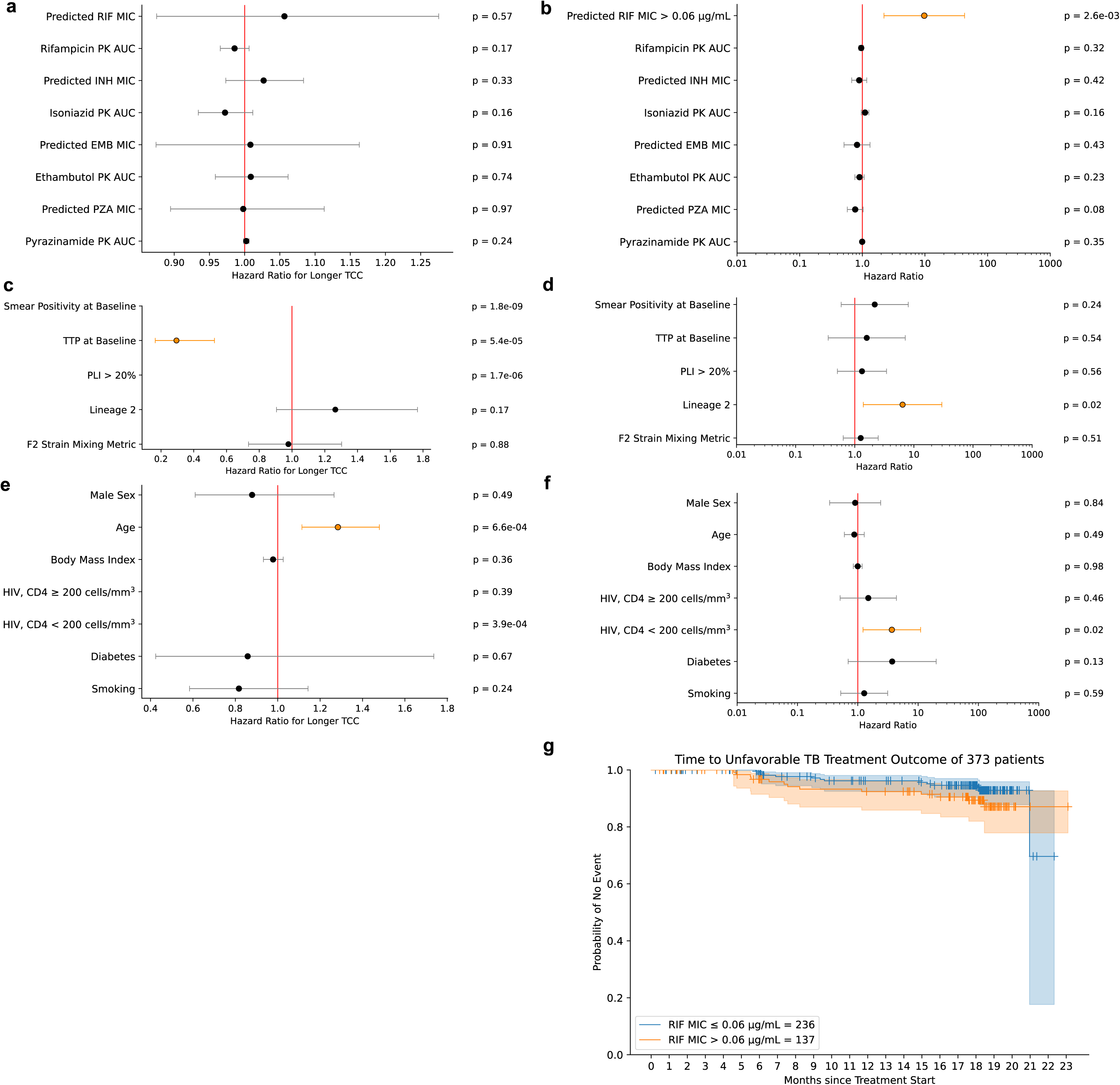
Associations between covariates and time to culture conversion (TCC) and unfavorable outcomes in multivariate models. Forest plots of associations between covariates and TCC **(a, c, e)** or unfavorable outcomes **(b, d, f)** in multivariate models with predicted MICs, predicted drug exposure, and patient characteristics**. g:** Kaplan-Meier estimate of event-free survival stratified by predicted rifampicin MIC (> 0.06 µg/mL or ≤ 0.06 µg/mL). Hazard ratios for continuous MIC variables are per increase in 0.1 µg/mL, and hazard ratios for age are per increase in 10 years. PK AUC: Pharmacokinetic (PK) area under curve (AUC) of drug concentration over time. These values were predicted from patient demographic data.^76^

Essential agreement rates between measured and predicted MICs for the four first-line drugs were 84%, 97%, 94%, and 96% for rifampicin, isoniazid, ethambutol, and pyrazinamide, respectively **(xtended Data Fig. 7e-h).** With the exception of isoniazid, there are few genetic differences in canonical resistance loci of these isolates **(Supplementary Results Section G)**.

### K. Predicted rifampicin MIC below the critical concentration is associated with TB treatment outcome

Using Cox proportional hazards regression, we studied both the time to sputum clearance of *Mtb* growth on culture (time to culture conversion, TCC) and a composite unfavorable final treatment outcome of 6-month treatment failure, death, or relapse **(Methods Section K, Extended Data Fig. 7a)**. We associated either CNN-predicted and phenotypically measured MICs for first-line drugs with outcome adjusting for patient confounders **(Table 4)** and predicted drug exposure. Measured pyrazinamide MICs were excluded as they were only available for 262 patients. Of the 373 participants included in the analysis, 27 patients (7%) had an unfavorable outcome (12 treatment failures, 3 deaths, and 12 relapses). Of the 353 participants with valid TCCs **(Methods Section K)**, the median TCC was 9 weeks, and 25% had *Mtb*-positive cultures at 12 weeks post enrollment **(Extended Data Fig. 7c).**

**Table 4:** Patient confounding variables tested in Cox models with log-rank test p-values of categorical variables based on Kaplan-Meier estimates of time to culture conversion. For the HIV+ variables, the log-rank test was calculated against the HIV– group. The continuous variables tested in the Cox model are listed in the table, but no log-rank test was performed because variables must be categorical. All variables were measured at baseline, within 2 weeks of starting TB treatment.

| Covariate | Variable Type | Log-Rank Test p-value in TCC KM Estimate |
| --- | --- | --- |
| Sex | Binary | 0.07 |
| Age | Continuous | NA |
| BMI | Continuous | NA |
| Smoking | Binary | 0.86 |
| HIV positivity and CD4 cell count | Categorical, 3 groups:<br>HIV–,<br>HIV+, < 200 CD4+ cells/mm<sup>3</sup>,<br>HIV+, $\geq 200$ CD4+ cells/mm<sup>3</sup> | HIV+, < 200 CD4+ cells/mm<sup>3</sup>: 0.39<br>HIV+, $\geq 200$ CD4+ cells/mm<sup>3</sup>: $3.9 \times 10^{-4}$ |
| Diabetes | Binary | 0.39 |
| Smear Positivity | Binary | $1.8 \times 10^{-9}$ |
| Time to culture positivity (TTP) | Continuous, hours | NA |
| % Lung Involvement > 20% | Binary | $1.7 \times 10^{-6}$ |
| Lineage 2 | Binary | 0.01 |
| F2 Lineage Mixing Metric | Continuous, 0-0.5 | NA |

The Cox models identify recognized associations between TCC or unfavorable outcomes and HIV with low CD4 cell count, age, smear grade, and time to culture positivity (TTP) **(Fig. 5, Supplementary Results Section H)**. We also validate a recently developed severity marker, high percentage of lung involvement (PLI) on chest X-ray.^52^ Cavitation from chest X-ray was excluded because it was not recorded for 26 patients, but we note that PLI has been reported to be a better predictor of unfavorable TB treatment outcomes than cavitation.^52^ We also note that lineage 2 *Mtb* isolates, which comprised 49% of the dataset, were associated with worse composite outcomes than L1, L3, and L4 **(Fig. 5d)**. We binarized the predicted rifampicin MIC at the median value of 0.06 µg/mL in MGIT medium due to the small numerical range of values. Measured MICs do not associate with either outcome **(Extended Data Fig. 8d-e),** but the CNN predicted rifampicin MIC > 0.06 µg/mL is associated with unfavorable outcomes (p =0.003, **Fig. 5b).**

## Discussion

We built a suite of convolutional neural networks that predict quantitative resistance for *Mtb* with diagnostic-grade accuracy. The CNNs achieve error rates within one MIC doubling, an error comparable to the experimental error rate of measuring MIC, across 8 first- and second-line anti-tubercular drugs including the critical drug bedaquiline **(Results Section D)**. For binary resistance classification, the CNNs perform comparably to or better than the 2023 WHO catalog classification method, despite being trained on a fraction of the data. CNNs have better accuracy for ethambutol, a drug known for genetic determinants that have subtle effects on MICs. CNNs also offer better sensitivity for isoniazid and levofloxacin and higher specificity for ethionamide. CNNs achieve high essential agreement for predicting sub-critical concentration MICs for 4 drugs in a rifampicin-susceptible TB cohort of 373 patients in South Africa in which MIC distributions differ considerably from those used for training the CNNs **(Results Section K)**.

Most notably, we find an association between higher predicted rifampicin MIC below the critical concentration and unfavorable treatment outcomes, adjusted for patient confounders. Measured rifampicin MIC was not associated with outcomes, which may be due to noise in MIC testing, especially at low testing concentrations. Time to initial culture conversion has been reported to be a poor proxy of final treatment outcomes which may be why we detect an association with composite outcomes but not with TCC.^53,54^ The predicted MIC association is in agreement with previously reported results by Colangeli *et al.*, in which similar rifampicin concentrations of 0.016-0.125 µg/mL were studied.^23^ Unlike the work by Colangeli et *al*, all participants with isoniazid resistance in our cohort had a favorable outcome, and we did not find an association between isoniazid MIC and unfavorable outcomes. Rifampicin, however, is recognized to be a more important drug in affecting sterilization and lasting cure than isoniazid, and hence our cohort size may not have had sufficient power to study effects of isoniazid MIC.^55^ We note that although HIV+ participants with low CD4 cell count had a higher risk of unfavorable outcomes, which is well known in tuberculosis disease,^56,57^ these participants also have significantly faster time to sputum clearance. This is likely due to the significantly lower bacterial burden at baseline in these participants. Although the bacteria is cleared from sputum, it may persist longer in participants with uncontrolled HIV infection, leading to worse long-term outcomes.

Due to universal limitations in dataset size and sampling, as well as the complexity of biological phenotypes, we hypothesized that ML models benefit substantially from encoding as much biological information as possible. We show that CNNs cannot learn the different consequences of nonsense and missense mutations on protein function from available nucleotide data alone and must be explicitly shown how each mutation affects protein sequence and length. Adding protein sequences and lineage/ancestry information improved model performance and reduced spurious associations. We also show that the core CNN architecture, which allows modeling of higher order interactions between genetic variants, is needed to achieve high performance. Simpler linear regression models that consider variants as independent are consistently less accurate across the 8 drugs we study.

Until recently, the prediction accuracy of ML models in biology was viewed as being at odds with model interpretability and their ability to learn causal relationships in biology. Here we show that CNNs can combine both state-of-art accuracy and interpretability. CNNs provide evidence to classify many mutations that have been graded uncertain in the WHO *Mtb* resistance mutation catalog, such as those in low-level activators of ethionamide. The models also identify putative binding pockets in proteins without solved structures, despite not being trained on 3D structures of the proteins. This would be especially important for new antibiotics for which the exact mechanisms are not yet known. By training models on MICs, rather than binary resistance phenotypes, we show that the models can associate genetic variants with drug hypersusceptibility and subtle shifts in MIC.

The CNN models are not without limitations. The models are still limited by the adequacy of training data, for example due to low rates of resistance for bedaquiline the CNN sensitivity for binary resistance prediction is still low. The bedaquiline CNN is nevertheless state-of-the-art in comparison with the WHO catalogue and now offers the prediction of MIC and borderline mutations. CNNs may assume that mutations of different types that occur on the same codon have similar effects, such as the silent variant pncA_c.3G>A and the missense variant ethR_p.3Ala. Loss of the start codons of *pncA* and *ethR* are associated with pyrazinamide resistance and ethionamide susceptibility, respectively, and the models extrapolate this association to other mutations on the same codons. This problem would be alleviated with richer training data and additional priors that weigh mutations such as start codons differently. Ultimately, CNNs cannot learn the biological impact of a mutation unless it or a neighboring mutation in either nucleotide or protein space is associated with the correct phenotype. Encoding additional information on protein stability or drug binding free energy^44^ may support models to accurately generalize to unseen or very rare mutations, particularly amino acid substitution mutations.

In this work, we show that machine learning models can reliably and accurately predict quantitative antibiotic resistance for *Mtb* with clinical-grade accuracy, and we verify that they learn from relevant genetic variants rather than spurious genetic correlations, setting them apart from previous efforts to predict MIC in *Mtb* **(Results Section E).**^58^ We find a significant association between higher rifampicin MIC and unfavorable treatment outcomes among individuals with rifampicin-susceptible *Mtb* infections, suggesting that low-level MICs are clinically relevant. There is increasing adoption of targeted sequencing for mutation detection for *Mtb* resistance diagnosis, and current commercial assays detect mutations in the majority of loci on which the CNN are trained (the CNNs were trained on additional loci to make use of information in non-canonical loci).^59^ CNNs could be deployed in the clinic, along with adequate information on interpretability for clinicians, to provide more comprehensive resistance information to inform treatment and dosing decisions.^60–62^ We speculate that the use of these models may now realize the full potential of precision treatment of tuberculosis as they achieve state-of-the-art accuracy for high level resistance and for the first time allow for the detection of borderline resistance that has been long elusive due to limited access to MICs. Future work should explore the use of the CNNs in clinical trials either prospectively or in retrospective reanalysis to study associations with outcome on larger datasets. Further, MIC prediction on the new and repurposed drugs – linezolid, delamanid, and pretomanid – may require different model architectures due to the current extreme rarity of resistant clinical isolates available for training.

## Methods

### A. Phenotypic Data

*Mtb* isolates were obtained from the Comprehensive Resistance Prediction for Tuberculosis: An International Consortium (CRyPTIC) project,^25^ the National Center for Biotechnology Information database, PATRIC,^26^ and the MIC-ML Consortium **(Supplementary Data 1)**. We trained single-drug models on MICs for eight anti-tubercular drugs – rifampicin, isoniazid, ethambutol, pyrazinamide, ethionamide, levofloxacin, moxifloxacin, and bedaquiline **(Table 1, Extended Data Fig. 1)**.

When measuring microbial MICs, specific concentrations of each drug are tested by different laboratories (typically 2-fold dilutions), making MIC a semi-continuous variable. If an isolate grows at a concentration of N, but not at 2N, the MIC is recorded as the range (N, 2N]. Because the true MIC within this interval is unknown, we built a custom loss function for the CNN **(Fig. 1b, Extended Data Fig. 2)**. The binned loss function computes mean absolute error (MAE) only for MIC predictions that lie outside the concentration interval associated with an isolate. To harmonize phenotypic data tested from different laboratories across testing media, we normalized all MICs to 7H10 medium using the ratio of the critical concentrations in the test medium and 7H10 medium, according to the method in Farhat *et al.* 2019.^4^ For pyrazinamide, all MICs were kept in MGIT medium as it was the testing medium for all available isolates, and for bedaquiline, all MICs were normalized to 7H11 medium.

For *Mtb*, resistance-conferring mutations arise primarily on the chromosome with no contribution from plasmids and horizontal gene transfer as in other bacteria. Instead of passing the entire *Mtb* genome into the models, we selected regions of interest for each drug to reduce computational complexity and overfitting to genetic variants that are difficult to interpret, and aid in targeted sequence-based efforts of diagnosing drug resistance in *Mtb*.^62^ Models were trained on genomic regions known or hypothesized to be relevant to resistance to the 8 drugs, including upstream promoter regions and intergenic regions between contiguous genes **(Table 1, Supplementary Table 1).**^7^ The following quality control steps were performed on a per-drug basis. We excluded isolates with a Group 1 catalog resistance mutation^16^ passing the quality control thresholds in **Methods Section C** and an MIC upper bound less than half of the critical concentration due to potential phenotypic mislabeling. We additionally removed isolates whose MIC range spans the drug’s critical concentration from model performance comparisons due to the inability to know if these isolates are drug-resistant or drug-susceptible from training, but we obtained post-training predictions for them. Isolate IDs, lineages, and MICs are in the **Supplement**. For all analyses, MICs were log2-transformed **(Extended Data Fig. 1)**. Isolates were split into training (80%) and testing (20%) sets, stratified by binary resistance phenotype and primary lineage. The testing set was further split into two groups of equal size, stratified by binary resistance phenotype: a validation set to determine early stopping for CNNs and the regularization parameter for regression models, and a hold-out test set on which to compute final model metrics. The MIC distributions for all drugs (without augmentation) are not significantly different between the combined train + validation set and the test set by a two-sided Kolmogorov-Smirnov test (p-values ≥ 0.99) (**Extended Data Fig. 1**).

### B. Phenotypic augmentation for bedaquiline and pyrazinamide

Due to the low number of bedaquiline-resistant isolates, leading to several resistance-conferring variants being absent from the training data **(Supplementary Results Section B)**, we augmented the training dataset with 827 bedaquiline-resistant isolates with high-quality binary phenotypes published by the 2nd edition of the WHO mutation catalog.^16^ These isolates were given an MIC of >0.25 µg/mL.

Due to the low number of total isolates with pyrazinamide MICs, we trained a separate CNN on binary pyrazinamide resistance phenotypes using the 1,021 isolates with MICs, which were binarized at the critical concentration of 100 µg/mL, and 13,086 additional publicly available isolates with high-quality binary phenotypes. This dataset has 20.0% resistant isolates, and the lineage distribution is more similar to those of the other drug datasets **(Extended Data Fig. 4a)**.

### C. Variant Calling Pipeline

Variant calling was performed as follows, with all software installed using conda (v23.10.0). FASTQ files were trimmed with fastp (v0.22.0), dropping trimmed reads shorter than 50 base pairs. Only reads mapping to a custom *Mtb* database built using 162 *Mtb* genomes with kraken (v2.1.2) were aligned with bwa-mem2 (v2.2.1) to the *Mycobacterium tuberculosis* H37Rv strain complete genome (NCBI RefSeq: NC_000962.3). The custom kraken database is composed of the H37Rv, F11, CTRI-2, and CDC1551 *Mtb* reference genomes and 151 *Mtb* short-read/long-read hybrid assemblies.^63^ After alignment, WGS runs were included if they had a median read depth ≥ 15 and at least 95% of the reference genome covered at a depth of 20x. For samples with multiple available sequencing runs, BAM files passing this alignment criteria were merged before variant calling. Duplicate reads were removed with picard (v2.20.4), followed by variant calling with pilon (v1.24). We excluded isolates with putative *Mtb* strain mixing as measured by an F2 metric^64^ greater than 0.1.

Additional variant filtering and annotation was done using bcftools (v1.9), samtools (v1.6), and snpEff (v5.2) using the same H37Rv reference sequence (GenBank assembly GCF_000195955.2_ASM19595v2) and the bacterial genetic code. We performed lineage typing using the fast-lineage-caller tool and used the lineages assigned by the Coll 2014 scheme.^30^

### D. Featurizing Nucleotide Sequences

As input for all models, we passed in a one-hot encoded nucleotide sequence matrix composed of all relevant loci for each drug. A single locus may contain many contiguous genes and their intergenic regions. Non-contiguous loci were passed in as separate channels. Multiple sequence alignments (MSAs) were generated for each contiguous locus using a custom Python script that inserts SNPs and indels from VCF files into the H37Rv reference sequence. Imprecise structural variants (pilon INFO/IMPRECISE = True) were excluded because they were not accurately resolved by the variant caller. We encoded variants with an alternative allele fraction (AF) > 0.75 as the alternative allele and variants with AF ≤ 0.25 as the reference allele. Low-quality SNVs, MNVs, and inframe indels (pilon FILTER ≠ PASS or Amb, pilon QUAL ≤ 10, read depth < 5, base quality < 20, mapping quality < 30, 0.25 < AF ≤ 0.75) were inserted as “N” nucleotides to distinguish them from variants of high confidence. However, we did not encode frameshift indels as missing because the insertion of “N” nucleotides can lead to improper translation to protein sequences. Frameshifts that passed all the QC criteria but had AF E (0.25, 0.75] were encoded as the alternative allele, and frameshifts that did not pass these QC criteria were left as reference.

The sequence alignment was one-hot encoded into a 5 x L matrix, where L is the length of the aligned locus. The 5 positions represent A, C, G, T, and the gap character, in this order. A = (1,0,0,0,0), C = (0,1,0,0,0), G = (0,0,1,0,0), T = (0,0,0,1,0), ‘-’ = (0,0,0,0,1), and N = (0,0,0,0,0). When there are multiple loci, each locus is a separate channel so that non-contiguous genes are never convolved over together. Shorter loci are padded at the end with gap characters. The final one-hot encoded input matrix for a single CNN sample is 5 x M x P, where M is the longest locus in the set and P is the number of loci. To feed data into regression models, the one-hot encoded matrix for each locus was flattened into a vector of length 5 x L_i_, where L_i_ is the length of the i^th^ locus. Vectors for all loci in the set were then concatenated, giving a full length of 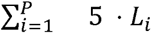. Because they do not contribute to the signal, features with no variation across the training set were removed for computational efficiency.

### E. Featurizing Amino Acid Sequences

We encoded amino acid biochemical properties, which implicitly encode protein lengths, to add additional biologically relevant information to the models and passed them into an additional convolutional block in the model, with the same layer structure as the nucleotide convolutional block **(Fig. 1a)**. We translated the nucleotide sequences in the MSAs and inspired by EVEscape,^27^ encoded three features – molecular weight (g/mol), isoelectric point, and hydrophobicity (Eisenberg scale) – as a 3 x K matrix, where K is the length of a given protein. Individual proteins were encoded as separate channels, even if they are translated from the same locus (*i.e.,* rpoB and rpoC proteins are in two separate channels in the amino acid block, while the corresponding gene sequences are concatenated into a single contiguous locus in the nucleotide block). Protein sequences for different isolates were left-aligned (*i.e.,* not a multiple sequence alignment), and then padded to the same length in the same manner as for the nucleotide sequences.

### F. Lineage SNP Inputs

To test the effect of including *Mtb* lineage information on model performance, we created a binary vector (1 = SNP present, 0 = absent or ambiguous) of 62 lineage SNPs based on the Coll *et al.,* 2014 SNP scheme for each isolate.^30^ The SNPs had to pass the same quality control thresholds previously described for the nucleotide MSA to be considered present. SNPs were concatenated with the flattened output of the convolutions and then passed through the two densely connected layers **(Fig. 1a)**. The distribution of primary lineage designations across all eight single-drug datasets is given in **Fig. 1c**. Because lineage SNPs do not significantly worsen performance, we included them in all drug models for downstream analyses to reduce overfitting to low-frequency genetic variants.^29^

### G. Model Design and Training

Nucleotide sequences and amino acid feature matrices were passed into two separate convolutional blocks, each with the same model architecture, as previously described:^7^ a shallow network with two sets of two convolutional layers and a max pooling layer. The outputs of the two convolutional blocks were flattened, concatenated together with lineage SNPs, and passed through two densely connected layers of 256 nodes each to obtain a final MIC prediction **(Fig. 1c)**. All layers except the output layer have ReLU activation.

Owing to the labor-intensive nature of measuring microbial MICs, specific concentrations of each drug are tested by different laboratories (typically 2-fold dilutions), making MIC a semi-continuous variable. If an isolate grows at a concentration of N, but not at 2N, the MIC is recorded as the range (N, 2N]. Because the true MIC within this interval is unknown, we built a custom loss function for the CNN **(Fig. 1b)**. The binned loss function only penalizes MIC predictions if they lie outside the concentration interval associated with an isolate. For predictions within the bounds, the error = 0. For predictions outside of these bounds, the loss is computed relative to the closer bound (i.e. the absolute error for a prediction of 0.7 for an isolate with a true MIC in the range (0.2, 0.4] would be 0.3). Our dataset is composed of MICs measured from different labs, which right-censor MICs differently: two isolates with the same true MIC of 100 µg/mL may be recorded as >2 µg/mL and >50 µg/mL because one laboratory did not test higher concentrations. The binned loss function accounts for these differences because it penalizes the predictions only if they are below the lower bounds of 2 and 50. We selected to minimize the binned mean absolute error because it is more robust to outliers than mean squared error or root mean squared error, especially given that the MIC datasets are considerably smaller than typical machine learning datasets. Additionally, squared error loss functions tend to weight larger mispredictions more than smaller mispredictions, and we wanted to minimize even small mispredictions of small MIC values.

The simplest and most interpretable model for predicting an unbounded continuous value is linear regression. Although regression models have achieved considerable success identifying genetic mutations that are associated with antibiotic resistance in *Mtb*,^4,34,65–68^ regression considers all features to be independent of one another and cannot learn from the locations of and distances between features. Further, the target must be a linear combination of inputs, which is not always the case for complex phenotypes driven by interactions between alleles (*i.e.* epistasis). Regression models were fit using scikit-learn’s (v1.4.2) Ridge regression function. We selected L2 regularization to correct for collinearity between nucleotides due to linkage disequilibrium in bacteria. The strength of the regularization parameter was selected among all powers of 10 from 10^-5^ to 10^5^ to minimize the validation loss. Because the scikit-learn implementation does not allow passing in multiple targets for each sample, it was trained to minimize mean absolute error using the midpoints of each MIC range.

We used 5-fold cross-validation, stratified by binary resistance phenotype, to compare model performance metrics on the hold-out test set. The cross-validated CNNs were trained using early stopping, until the test loss did not decrease by at least 1% for 200 (*i.e.,* the patience period is 200 epochs long). Significance testing was performed using Welch’s t-test between the 5 splits of two given models. We compared binned mean absolute error (referred to simply as “mean absolute error”) between models. For sensitivity and specificity, 95% confidence interval bounds were computed on the cross-validation results as 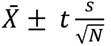, where N = 5, the number of cross-validation folds; 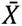 and s are the mean and standard deviation, respectively, of the five folds; and t is the score from the Student’s t-distribution with α = 0.05 and degrees of freedom = N - 1 = 4. After cross-validation, a final model was trained on the full dataset for each drug using these same training parameters for downstream analyses.

For the binary pyrazinamide models, all training parameters, including the number of training epochs and stopping criterion, were the same as for the quantitative models, but the models were trained using binary cross-entropy as the loss. We selected the threshold for binarizing the predicted scores to maximize sensitivity and specificity.

### H. Saliency Mapping

We used the DeepLIFT attribution algorithm^69^ as implemented in DeepExplain^70^ to measure how strongly each categorical feature (nucleotide identity and lineage SNP presence vs. absence) affects model predictions. By comparing each sample to a reference sequence (H37Rv), the algorithm assigns a “saliency score” to each feature that differs from the reference in each training isolate. The score for a single nucleotide position is the sum of scores across the four values associated with the four non-reference alleles at that site, and we computed an aggregate maximum (positive) and minimum (negative) score for each position. Lineage saliency scores are similarly computed with the reference lineage type being the reference strain H37Rv (i.e. the vector of lineage SNPs composed entirely of 0s). We performed a permutation test to assess the significance of the scores. The phenotype labels for each dataset were randomly shuffled, and a new model was trained using the same previously selected regularization parameter until the loss did not decrease by 1% for 100 epochs. This process was repeated 10 times. For positions with a non-zero maximum or minimum saliency score, p-values were computed using the permuted scores. For sites with a 0 score, there is no meaningful p-value because the permuted scores are also zero.

We computed correlations between the saliency scores and the average log2-MIC observed in the training dataset for different lineage SNPs. We included only sublineages with at least 2 isolates and with significant saliency scores according to the above permutation test.

### I. *In silico* mutagenesis

To isolate the effects of individual genetic variants on predicted MIC, we performed *in silico* mutagenesis experiments. Individual mutations or lineage SNPs were inserted into the H37Rv reference to observe how the trained CNNs predict MIC on a sequence with a single genetic change. Because the CNN can only make new predictions for input matrices of the same dimensions (longest locus length x one-hot encodings x number of loci) as the data it was trained on, insertions at positions where there were no insertions in the training dataset could not be tested because they increase the size of the multiple sequence alignment. Due to some naming inconsistencies in the WHO mutation catalog, we derived the nucleotide change using the name of a mutation according to the Sequence Ontology nomenclature used in the catalog and verified the accuracy by annotating with snpEff (v5.2). As in the catalog, we considered loss of function (LoF) mutations for coding regions to be loss of start codons, early stop codons, frameshift mutations, and gene deletions / ablations. For amino acids encoded by multiple codons, the codon with the smallest edit distance from the reference codon was selected to encode it in the *in silico* sequence. Frameshift variants were not included in this analysis because different nucleotide changes of drastically different sizes could all result in the same frameshift mutation in the WHO catalog naming system.

We inserted the 62 Coll lineage SNPs and all compatible mutations from the 2^nd^ edition of the WHO resistance mutation catalog into H37Rv. We additionally performed site-saturation mutagenesis for some proteins by mutating each residue to the other 19 possible amino acids and the stop codon. For the first amino acid, all changes except to the six alternative start codons in bacteria were considered to be start lost mutations. Because all changes to the first codon are either loss of start mutations or initiator codon mutations that are translated to methionine, the first codon was excluded from the clustering computation because it does not have missense mutations. The *in silico* mutagenesis procedures yield a predicted MIC for H37Rv. We used this predicted value to compute fold changes for mutations.

### J. 3D Space Clustering Computation

The Getis-Ord statistic is used to measure spatial association of a user-defined metric and identify hot and cold spots ^40^. For the i^th^ object in a group, the Getis-Ord (G_i_*) statistic is the z-score of the metric, weighted by the distance between the object and all other objects. It therefore estimates whether the object has a very high (positive) or low (negative) z-score and is also surrounded by objects with extreme z-scores. We computed G_i_* statistics using the average log fold change of predicted MIC (log_2_FC) across the 19 possible missense mutations at each residue of a protein, excluding start lost mutations.

A permutation test was then performed with 10,000 replicates to determine in what fraction of permuted structures the i^th^ residue has a G_i_* statistic at least as extreme as the test score. FDR correction was performed using the Benjamini-Hochberg method, and we selected an FDR q-value of 0.05 as the threshold for significant clustering. We considered the i^th^ residue to be a resistance-associated “hotspot” if G_i_* > 0 and a susceptibility-associated “coldspot” if G_i_* < 0 and log_2_FC_i_ < 0. A residue with a significant Getis-Ord statistic (Benjamini-Hochberg-corrected p-value ≤ 0.05) implies it is surrounded by residues with predicted MICs significantly higher or significantly lower than the average for the entire protein. A residue with a high log_2_FC may not necessarily have a significant G_i_* statistic if it is not also surrounded by residues with high predicted MICs and vice versa for low log_2_FC.

The proteins for which we performed 3D clustering are as follows: rpoB (UniProt: P9WGY9, PDB: 5UHB, chain C), gyrA and gyrB (UniProt: P9WG45, P9WG47; PDB: 5BS8), pncA (UniProt: I6XD65, PDB: 3PL1), katG (UniProt: P9WIE5, PDB: 4C51), ethA (UniProt: P9WNF9), and Rv0678 (UniProt: I6Y8F7, PDB: 4NB5).

### K. Patient Outcomes Models

To test if MICs are associated with unfavorable TB treatment outcome, we built multivariate Cox proportional hazards models to associate clinical and microbiological data from participants in the TRUST study^51^ with two events – sputum culture conversion and unfavorable outcomes. Participants in this study have rifampicin-susceptible pulmonary TB diagnosed by GeneXpert and were all treated with a 6-month regimen of RIPE (rifampicin, isoniazid, pyrazinamide, and ethambutol), followed by 12 months of follow-up. Time zero for each participant is the time of initial screening and treatment initiation. A total of 440 individuals were enrolled at the time this analysis was conducted. 60% are male, 28% have HIV, 38% had a previous TB disease episode, 4% have diabetes, and 54% have a body mass index (BMI) < 18. A total of 373 participants remained after excluding participants who changed treatment regimens due to developing rifampicin resistance or needing a liver-friendly regimen, participants without WGS of their bacterial cultures performed within the first two weeks of enrollment, and participants with incomplete data **(Extended Data Fig. 7a)**. One participant did not have MICs measured, whom we excluded only from the models with measured MICs as covariates.

Unfavorable treatment outcomes were defined as treatment failure, TB disease relapse, or death. Treatment failure was assessed after 6 months of treatment, either by a positive culture or symptomatically. Relapse was determined if a participant was sputum- or smear-positive at the conclusion of the 12-month follow-up period. The median duration of enrollment of the 373 patients was 545 days, and the longest was 835 days. We censored 5 of the 8 patients whose deaths were attributed to non-TB causes, specifically cancer (N = 3), COVID-19 (N = 1), and accidental deaths (N = 1), leaving 3 TB-associated deaths. All other patients were right-censored at their study completion date or their last known clinic appointment if they were lost to follow-up (N = 35) or were still enrolled in the study (N = 12) at the time of analysis.

Sputum was sampled weekly for the first 12 weeks of treatment. Time to negative sputum culture conversion (TCC) was defined as the number of weeks between the time of treatment initiation and the first of two consecutive *Mtb*-negative sputum cultures not followed by another positive culture. To be included in the TCC analysis, participants had to have at least 3 uncontaminated sputum samples and a positive *Mtb* culture within the first 5 weeks of initial screening, leaving 353/373 patients remaining for the TCC analysis. We imputed missing cultures after this using the mice package (3.16.0) in R (4.4.2).^71^ To impute each culture, we used age, sex, BMI, HIV/CD4 status, smoked substance use (methamphetamine, methaqualone, or cannabis), alcohol use, isoniazid resistance, time to culture positivity at baseline, and all smear positivity and culture results taken before and at the same week as the culture to be imputed, similar to as done previously.^71^ TCC was computed on each of 30 imputed datasets, and model results were pooled across 30 imputations using Rubin’s rules.^72^

We selected 11 relevant clinical covariates using domain knowledge and past evidence for association with poor TB treatment outcomes **(Table 4).**^73–75^ Percent lung involvement and drug exposure measured by the area under the pharmacokinetic ROC curve^76^ were predicted from chest x-ray images and patient metadata, respectively, while the other independent variables were determined from patient health records. Based on past work with this and other data,^52^ we binarized percent lung involvement at 20% to generate a variable “high lung involvement.” Cavitation was excluded due to a high rate of missingness. We additionally added a binary variable for *Mtb* lineage (L2 vs. all other lineages due to the low prevalence of L1 and L3 isolates) and included the F2 strain mixing metric at baseline.^64^

To satisfy the Cox proportional hazards assumption, we stratified by smear positivity, high lung involvement, and HIV/CD4 status in the TCC model only **(Fig. 5d-f)**. The proportional hazards assumption (p > 0.01, Schoenfeld residuals test) was met for all covariates in the unfavorable outcomes model. To these models, we added measured or CNN-predicted MICs (log_2_-transformed) for the four first-line drugs to determine if the MICs were significantly associated with outcomes, conditioned on the presence of other relevant patient characteristics. We also controlled for predicted drug exposure in each model with MICs.^76^

## Data Availability

All data produced are available online at https://github.com/sanju99/MtbQuantCNN, with the exception of the unpublished human subject data, which is available upon reasonable request to the authors.

## Supplement

## Results

### A. Amino acid biochemical properties are required for CNN accuracy and correct attribution of resistance to genetic features

As part of the *in silico* validation using WHO catalog mutations, we observed that nucleotide-only CNNs failed to learn that nonsense mutations in non-essential genes are associated with resistance. We demonstrate this with *pncA*, which encodes the enzyme pyrazinamidase (PZase) that activates pyrazinamide to its active form pyrazinoic acid.^79^ Loss of function (LoF) mutations in pncA lead to pyrazinamide resistance because the drug remains inactive, and therefore, all LoF mutations are predicted to cause pyrazinamide resistance in the WHO catalog.^16^ However, we observed that only 13/186 (7%) nonsense mutations along the length of the pncA protein were predicted to elevate MIC above the critical concentration of 100 µg/mL **(Extended Data Fig. 3a)**. We reasoned that the model failed to make accurate predictions for nonsense mutations because it did not observe the corresponding change to the amino acid sequence. Inspired by EVEscape,^27^ we therefore encoded three features – molecular weight (g/mol), isoelectric point, and hydrophobicity (Eisenberg scale) for each protein encoded by the genes passed into the nucleotide convolutional block **(Fig. 1b).** This information implicitly encodes the translated protein length. All models in this work include the amino acid properties.

Without the amino acid biophysical properties, the pyrazinamide quantitative CNN predicted similar MICs for nonsense mutations in *pncA* and missense mutations that are graded Uncertain in the WHO mutation catalog at the same codons **(Extended Data Fig. 3b)**. Among variants found in the catalog, the CNN with amino acid biophysical properties predicts substantially higher MICs for nonsense mutations than for corresponding missense mutations (averaged across the observed missense mutations) on the same codons (p << 0.01, one-sided paired t-test) **(Extended Data Fig. 3d)**, while the CNN without amino acid biophysical properties does not **(Extended Data Fig. 3b)** (p = 0.74, one-sided paired t-test). Additionally, the pyrazinamide CNN with amino acid properties has significantly lower MAE than the model without (ΔΜΑΕ = 0.33 log2-MIC units, p = 0.03, one-sided Welch’s t-test) **(Extended Data Fig. 3e)**.

Although only a subset of LoF variants are in the training data, these models are able to accurately extrapolate resistance effects to other loss of function (LoF) mutations presented to the model during *in silico* mutagenesis in line with the 2023 WHO mutation catalog recommendation^24^ **(Extended Data Fig. 3c)** to consider nearly all *pncA* nonsense variants causal of pyrazinamide resistance. The only pncA residues at which nonsense mutations are not predicted to cause pyrazinamide resistance occur after residue 175 of the 186 amino acid long protein. However, we note that the CNN is susceptible to confusion between different mutation types on the same codon. Nonsense mutations around residue 66 in *pncA* are predicted to cause a lower MIC than nonsense mutations elsewhere, which is likely due to confusion with the variant pncA_p.Ser66Leu, which is not associated with pyrazinamide resistance **(Extended Data Fig. 3c)**.

The CNN’s inability to learn the effects of nonsense mutations from nucleotide sequences only is likely because in the nucleotide feature space, nonsense, missense, and synonymous mutations cause local changes, affecting only 1-3 nucleotides each, but when translated to amino acid feature space, the sequence changes are much more pronounced for nonsense mutations than the other types. Nonsense mutations are rare in the training datasets for drugs that must be activated by *Mtb* enzymes – 13, 18, and 158 isolates have a nonsense mutation in *pncA, katG,* and *ethA,* the activators of pyrazinamide, isoniazid, and ethionamide, respectively. Therefore, with these small numbers, CNNs can not learn the effects of nonsense mutations from nucleotide sequences alone.

### B. Augmentation of bedaquiline dataset improves resistance variant detection

The unaugmented bedaquiline model was unable to determine that the two missense “1) Assoc w R variants” in the WHO *Mtb* resistance mutation catalog were associated with bedaquiline resistance **(Extended Data Fig. 4d)** because it had only observed them in 5 isolates, only one of which has an MIC greater than the bedaquiline critical concentration. After augmentation, which increased the number of resistance variants in the training data, we observe a significant increase in the *in silico*-predicted MICs for Group 1 and 2 resistance variants from the catalog **(Extended Data Fig. 4d).** Several uncertain mutations in the catalog also were newly predicted to be associated with resistance. The augmented model does not have significantly lower MAE than the unaugmented model (p = 0.53, one-sided Welch’s t-test, **Extended Data Fig. 4e)**. Additionally, we confirmed that the augmented model does not systematically predict higher MICs for all test-set isolates. The predictions are not significantly larger for the augmented model (p = 0.93, Wilcoxon signed-rank test **Extended Data Fig. 4f**).

### C. Comparison between CNN essential agreement and published state-of-the-art models

The essential agreement rates of the CNNs, when restricted to the CRyPTIC data, are on average 0.85 percentage points lower than those reported by the CRyPTIC consortium using XGBoost models trained on k-mers extracted from raw sequencing reads.^58^ However, the most salient k-mers used to train the XGBoost models are frequently not in genes with causal effects on resistance. For example, the top hits for ethambutol are in *katG*, likely due to the high probability that an ethambutol-resistant sample is also isoniazid-resistant. Therefore, the XGBoost models are learning to make predictions based on correlation between resistances to different drugs, rather than from causal genetic variants.^7^

### D. Lineage Saliency Scores associate with lineage distribution of MICs

We studied the saliency (*i.e.,* importance) of the 62 lineage SNP barcode using the DeepLIFT algorithm^69^ based on a comparison to a reference barcode with none of the lineage SNPs present. This approach measures how much the MIC prediction increases with the presence of each mutation.

50-58 of the 62 lineage SNPs have a significant saliency score and are present in at least 2 training isolates across the 7 drug models, excluding pyrazinamide because the lineage barcode worsened model performance. For five drugs, all of which had an improvement in MAE with the lineage barcode, we observe significant Spearman correlations between these significant saliency scores and the average MICs of the associated sublineages in the training data: bedaquiline (0.28, p = 0.05), ethambutol (ρ = 0.35, p = 0.01), ethionamide = (0.31, p = 0.03), levofloxacin (0.36, p = 0.01), and moxifloxacin (0.48, p = 0) **(Extended Data Fig. 9)**. The remaining insignificant associations are for rifampicin (ρ = 0.06, p = 0.65) and isoniazid (0.24, p = 0.07) **(Extended Data Fig. 9)**. Differential drug susceptibility across the *Mtb* lineages has been reported,^80^ and further study is needed to confirm the associations found here.

Further, for synthetic sequences with individual lineage SNPs introduced into H37Rv, single lineage SNPs should not be predicted to cause resistance. We observe predicted MICs less than the critical concentration for all drugs except pyrazinamide **(Extended Data Fig. 9)**. This is consistent with the lineage barcode worsening performance for pyrazinamide, likely due to severe skew in the lineage distribution.

### E. Discrepancies between CNN predictions and the WHO catalogue as interrogated through *in silico* mutagenesis

We note that across the 8 quantitative drug models presented here, only two silent mutations from the 2^nd^ version of the WHO mutation catalog^24^ are predicted to cause resistance – pncA_c.3G>A (initiator codon variant) for pyrazinamide and mmpL5_c.2889G>A for bedaquiline. pncA_c.3G>A is absent from the training dataset, and in fact, the only variant in the training set that overlaps the start codon is an 845 base pair deletion that ablates the start codon (loss of the *pncA* start codon is known to cause pyrazinamide resistance). Due to the limited training set size, the CNN wrongly learned to associate any changes on the first codon with resistance.

mmpL5_c.2889G>A is found in 7 bedaquiline-resistant isolates in the training dataset without a known resistance variant. All are L2.2.1 and are isolates with binary data used to augment the bedaquiline dataset, and it was not predicted to elevate resistance above the CC in the unaugmented model **(Extended Data Fig. 4d)**. Lineage confounders or novel causes of resistance may be the cause of this misclassification. Across the 8 drugs, a total of 82 Group 1 variants in the WHO mutation catalog are not predicted to be associated with MICs greater than the respective critical concentrations (CC). 67 of these variants are related to pyrazinamide resistance due to the very small size of the MIC training dataset, but the binary model correctly identifies 119/124 Group 1 pyrazinamide resistance mutations. Both mutations in Group 1 for bedaquiline were predicted to associate with MICs above the CC by the augmented model, and for the remaining six drugs, 1-6 mutations for each drug are missed **(Extended Data Fig. 10)**. The four missed Group 1 variants for rifampicin and moxifloxacin all have elevated predicted MICs and are classified as borderline or intermediate resistance variants. For isoniazid, the missed variant – katG_p.Trp328Leu – is found in a single training isolate, which is likely insufficient for the model to learn it.

### F. CNNs identify genetic associations with hypersusceptibility

Different from models of binary resistance, models trained on MICs may identify mutations associated with drug hypersusceptibility. We identify 23 non-silent mutations from the WHO catalog with predicted log_2_FC ≤ -1, suggesting association with hypersusceptibility **(Table 3).** The majority of these (57%) are LoF or inframe deletion variants in *mmpL5*, which are known to confer bedaquiline hypersusceptibility^16,81^. Two LoF mutations in *ethR* are associated with ethionamide hypersusceptibility, consistent with the known repressive activity of ethR on *ethA* transcription.^82^ Three missense mutations in *ethA* at residues 391, 393, and 397 are associated with lower ethionamide MIC, and this is also reflected in the site-saturation mutagenesis results **(Fig. 4h).** The lowest fold changes are for mutations from the hydrophobic amino acids leucine and tryptophan to the positively charged amino acids lysine and arginine **(Fig. 4h)**. These residues are also located near the predicted FAD-binding site in ethA, particularly tryptophan 391, whose side chain extends inwards towards the predicted FAD and is within 3.1 Å of it **(Fig. 4i)**. We hypothesize that changes to these residues near the likely active site of the enzyme increases activation of ethionamide.

### G. MIC predictions and genetic mutations for participants in the TRUST cohort

Restricting to the 25 isoniazid-resistant participants, the essential agreement rate for isoniazid is 84%. Thirteen participants have measured isoniazid resistance explained by canonical resistance mutations. Of these, 12 were predicted by the models to be isoniazid-resistant, and the missed participant has a low-frequency resistance variant due to a co-infection between lineages 2 and 4. All mutations in genes known to confer resistance can be found in **Supplementary Data 8.** No samples contain rifampicin resistance-associated variants. One sample contains a known ethambutol resistance variant (embB_p.Met306Val), and another contains a known pyrazinamide resistance variant (pncA_p.Cys14Arg).

### H. Significant associations between patient covariates and treatment outcomes

To satisfy the proportional hazards assumption, we stratified the TCC model by smear positivity at baseline, percent of lung involvement (PLI) > 20%, and HIV/CD4 status. Therefore, there are no hazard ratios reported for these variables. Instead, we report the results of the log-rank test **(Extended Data Fig. 8a-c, Table 4)**. The probabilities of sputum remaining *Mtb*-positive longer were significantly higher for participants with positive smear (p = 1.8 x 10^-9^), high PLI (p = 1.7 x 10^-6^), and HIV+ participants with CD4 < 200 cells/mm^3^ (p = 3.9 x 10^-4^). There is no significant difference between HIV– participants and HIV+ participants with CD4 ≥ 200 cells/mm^3^ by the log-rank test (p = 0.39) **(Table 4).** Age was significantly associated with longer TCC, time to culture positivity (TTP) with shorter TCC, and HIV positivity with low CD4 cell count with unfavorable outcomes **(Fig. 5)**.

---

**Supplementary Table 1.** Genetic loci used for training the models. Coordinates are with respect to H37Rv and were determined from Mycobrowser.^78^ Individual genes within a contiguous locus are delimited by hyphens.

| Locus Genes | Start | End | Sense |
| --- | --- | --- | --- |
| *gyrB-gyrA* | 4998 | 9818 | + |
| *mshA* | 575301 | 576790 | + |
| *Rv0565c* | 656010 | 657547 | – |
| *rpoB-rpoC* | 759479 | 767320 | + |
| *mmpL5-mmpS5* | 775586 | 778989 | – |
| *Rv0678* | 778906 | 779487 | + |
| *atpE* | 1460997 | 1461290 | + |
| *fabG1-inhA* | 1673300 | 1675011 | + |
| *rpsA* | 1833493 | 1834987 | + |
| *ndh* | 2101651 | 2103183 | – |
| *katG-furA* | 2153889 | 2156705 | – |
| *Rv2042c-pncA* | 2287884 | 2289281 | – |
| *fabD-acpM-kasA-kasB-accD6* | 2516549 | 2522164 | + |
| *oxyR'-ahpC-ahpD* | 2725571 | 2727339 | + |
| *pepQ* | 2859300 | 2860451 | – |
| *Rv3083* | 3448427 | 3449991 | + |
| *clpC1* | 4038158 | 4040759 | – |
| *panD-panC-Rv3603c* | 4043862 | 4046302 | – |
| *aftA-embC-embA-embB* | 4237930 | 4249810 | + |

| **Locus Genes** | **Start** | **End** | **Sense** |
| --- | --- | --- | --- |
| *gyrB-gyrA* | 4998 | 9818 | + |
| *mshA* | 575301 | 576790 | + |
| *Rv0565c* | 656010 | 657547 | – |
| *rpoB-rpoC* | 759479 | 767320 | + |
| *mmpL5-mmpS5* | 775586 | 778989 | – |
| *Rv0678* | 778906 | 779487 | + |
| *atpE* | 1460997 | 1461290 | + |
| *fabG1-inhA* | 1673300 | 1675011 | + |
| *rpsA* | 1833493 | 1834987 | + |
| *ndh* | 2101651 | 2103183 | – |
| *katG-furA* | 2153889 | 2156705 | – |
| *Rv2042c-pncA* | 2287884 | 2289281 | – |
| *aftB-ubiA-Rv3807c-glfT2-glf* | 4266953 | 4273738 | – |
| *ethA* | 4326004 | 4327548 | – |
| *ethR* | 4327474 | 4328199 | + |

**Supplementary Table 2.** Binary classification metrics of CNNs and WHO catalog classification on the hold-out test set. The CNN predictions were dichotomized at the critical concentrations. TN: true negatives, FP: false positives, FN: false negatives, TP: true positives. The confidence intervals are plotted in **Fig. 3a-b**, and the confidence intervals and the F1 scores can be found in **Supplementary Data 6.**

| **Drug** | **Model** | **TN** | **FP** | **FN** | **TP** | **Sensitivity** | **Specificity** |
| --- | --- | --- | --- | --- | --- | --- | --- |
| Bedaquiline | CNN | 1775 | 25 | 14 | 12 | 46.2 | 98.6 |
| Bedaquiline | Catalog | 1772 | 28 | 18 | 19 | 51.4 | 98.4 |
| Ethambutol | CNN | 1303 | 145 | 98 | 451 | 82.1 | 90.0 |
| Ethambutol | Catalog | 1206 | 242 | 245 | 574 | 70.1 | 83.3 |
| Ethionamide | CNN | 1614 | 73 | 132 | 205 | 60.8 | 95.7 |
| Ethionamide | Catalog | 1498 | 189 | 115 | 292 | 71.7 | 88.8 |
| Isoniazid | CNN | 1056 | 35 | 69 | 1254 | 94.8 | 96.8 |
| Isoniazid | Catalog | 1060 | 31 | 205 | 1281 | 86.2 | 97.2 |
| Levofloxacin | CNN | 1245 | 22 | 26 | 267 | 91.1 | 98.3 |
| Levofloxacin | Catalog | 1241 | 26 | 43 | 250 | 85.3 | 97.9 |
| Moxifloxacin | CNN | 1482 | 54 | 53 | 296 | 84.8 | 96.5 |
| Moxifloxacin | Catalog | 1477 | 59 | 62 | 289 | 82.3 | 96.2 |
| Pyrazinamide | CNN | 94 | 5 | 28 | 78 | 73.6 | 94.9 |
| Pyrazinamide | Catalog | 87 | 12 | 10 | 96 | 90.6 | 87.9 |
| Rifampicin | CNN | 1142 | 20 | 40 | 1104 | 96.5 | 98.3 |
| Rifampicin | Catalog | 1133 | 29 | 58 | 1149 | 95.2 | 97.5 |

## Extended Data

**Extended Data Figure 1.**
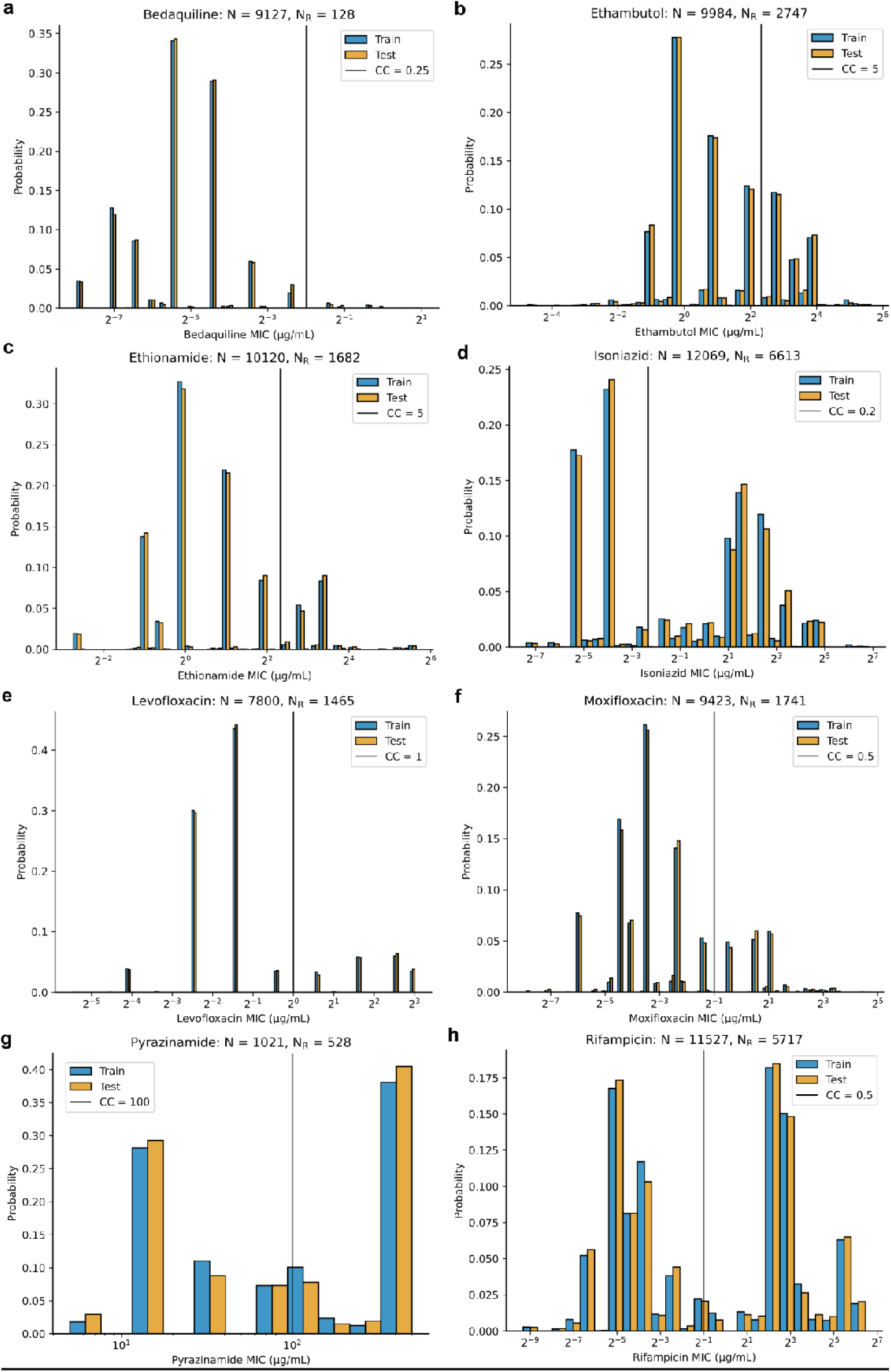
Histograms of measured MICs for 8 drugs, separated into training and testing splits. In order: bedaquiline **(a)**, ethambutol, ethionamide, isoniazid, levofloxacin, moxifloxacin, pyrazinamide, and rifampicin **(h).** The train (80%) and test (20%) datasets were split stratified by binary phenotype and primary *Mtb* lineage. The train and test distribution probabilities are normalized separately for viewing because the test set is considerably smaller. All MICs and critical concentrations (CCs) are in 7H10 media, except for pyrazinamide (MGIT). A two-sided Kolmogorov-Smirnov test was performed to compare the two MIC distributions for each drug, with p-values ≥ 0.99. All distributions shown are without augmentation.

**Extended Data Figure 2:**
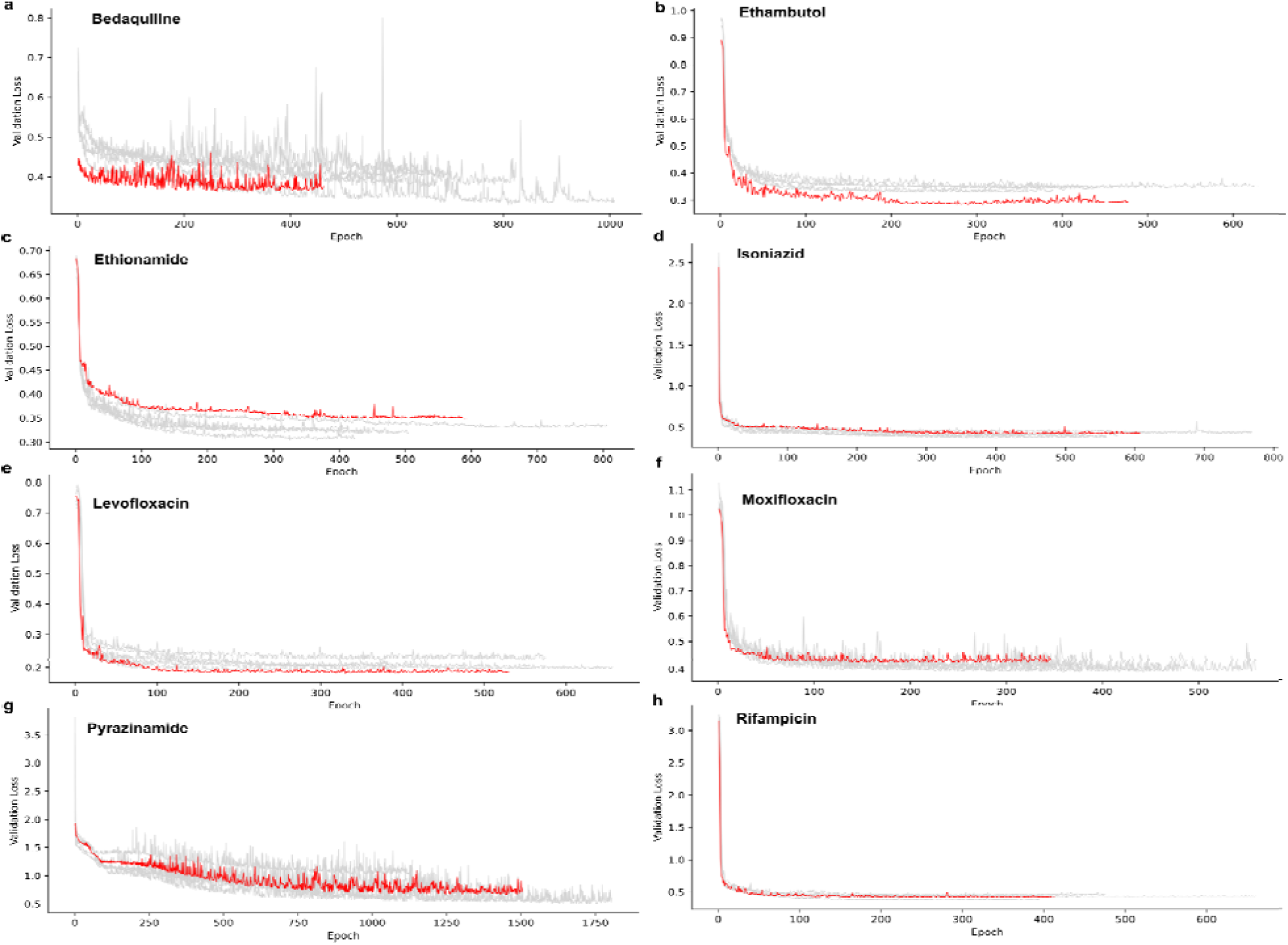
Loss curves for 8 single drug quantitative CNNs with lineage SNPs, except for pyrazinamide. In order: bedaquiline **(a)**, ethambutol, ethionamide, isoniazid, levofloxacin, moxifloxacin, pyrazinamide, and rifampicin **(h).** For each plot, there are five gray lines corresponding with the five splits during cross-validation. Each of these was trained on 80% of the training data, and the stopping criterion was determined using the remaining 20% of the training data, on which the plotted loss was determined, except for bedaquiline, which has a slightly larger training set due to augmentation. The red line is the loss on the test dataset while the model was trained on the full training dataset. The patience period of 200 epochs is included in all panels, which in many cases is evident by a slight increase in the loss. The gray lines are the models used to compare model performances, and the red line corresponds with the model used for all other downstream analyses.

**Extended Data Figure 3.**
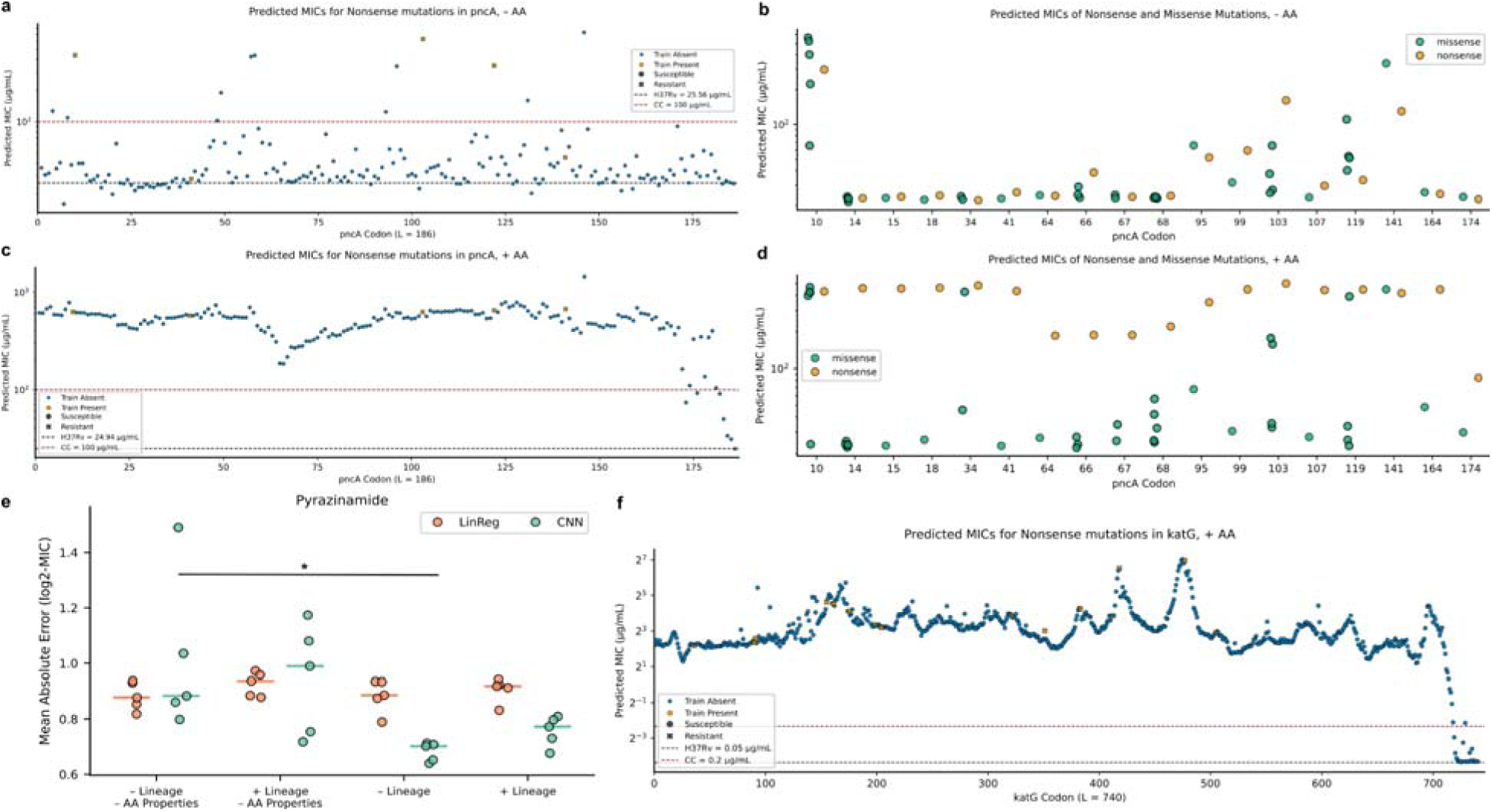
Encoding amino acid biophysical properties improves biological accuracy of the PZA quantitative CNN. *In silico* predicted MICs for missense and nonsense mutations from the 2023 WHO catalog on the same codon by PZA models without **(a)** and with **(c)** AA properties. Predicted MICs for nonsense mutations at every position in the pncA protein for models without **(b)** and with **(d)** AA properties. Nonsense mutations present in the training dataset are shown in orange and are slightly larger than the other points for readability. For nonsense mutations that are found in the training dataset, they are represented with an “X” marker if they have a measured resistant MIC (> 100 µg/mL). **e:** Comparison of MAE across pyrazinamide models with and without AA properties and with and without the lineage barcode. Inclusion of AA properties significantly reduces MAE for the model without the lineage barcode (p = 0.03, one-sided Welch’s t-test) and marginally for the model with the linear barcode (p = 0.053) **f:** Predicted MICs of synthetic nonsense mutations at each position in the katG monomer.

**Extended Data Figure 4.**
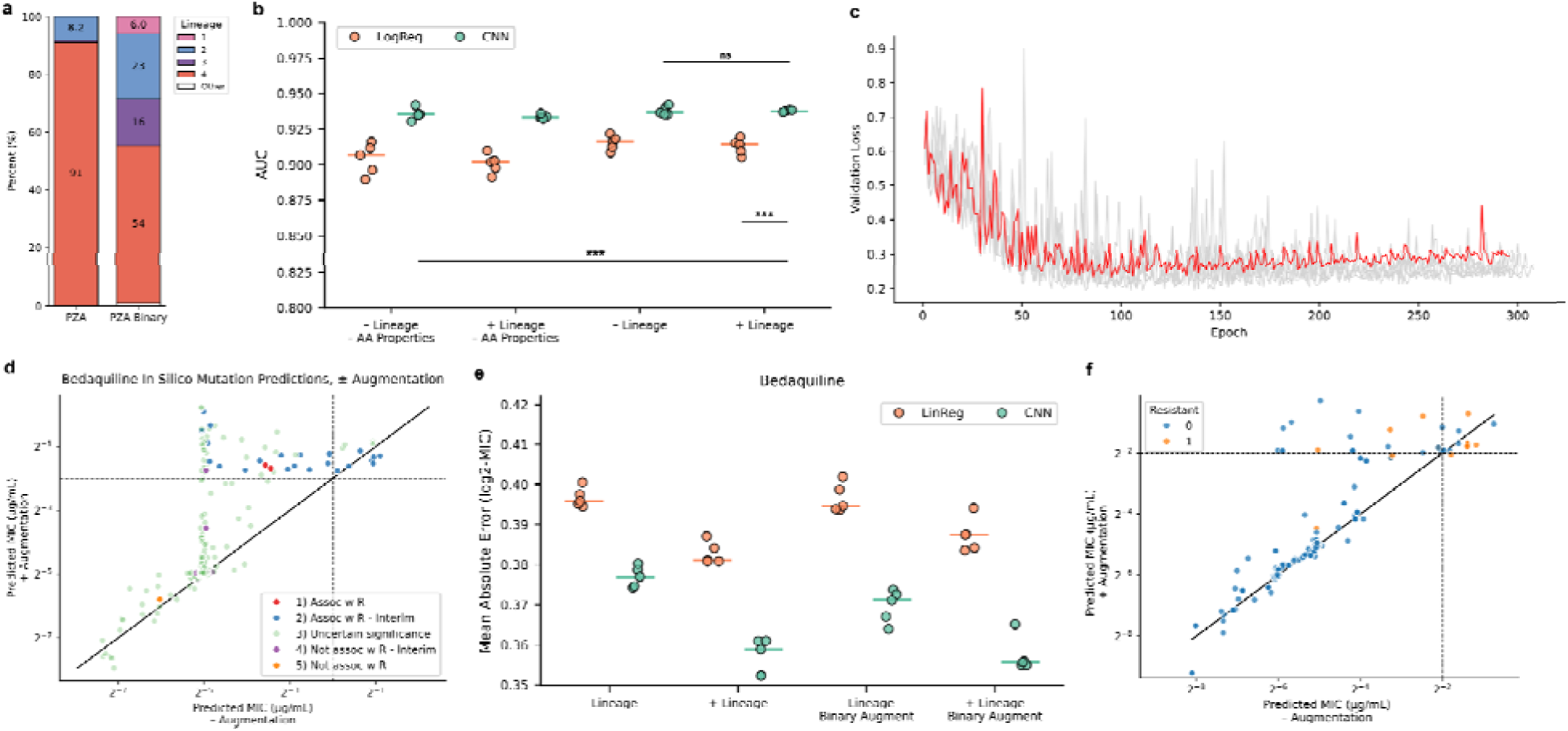
Augmented pyrazinamide and bedaquiline model results. **a:** Lineage comparison between pyrazinamide quantitative and binary datasets used in this work. **b:** AUC comparison between pyrazinamide binary CNNs and logistic regression models, with and without amino acid properties and with and without the lineage barcode. **c:** Loss (binary cross-entropy) curve for the pyrazinamide model with the lineage barcode and amino acid properties. The patience period of 200 epochs is included in the loss curve. **d:** *In silico* bedaquiline MIC predictions for 737 mutations from the WHO *Mtb* resistance mutation catalog between augmented (y-axis) and unaugmented (x-axis) models. **e:** Mean absolute error comparisons between augmented and unaugmented bedaquiline models, with and without the lineage barcode. **f:** Scatterplot of predictions on the test set isolates between augmented (y-axis) and unaugmented (x-axis) bedaquiline models. Dashed vertical and horizontal lines on panels **d** and **f** correspond with the critical concentration of 0.25 µg/mL for bedaquiline. Solid black lines on panels **d** and **f** are 45-degree lines.

**Extended Data Figure 5.**
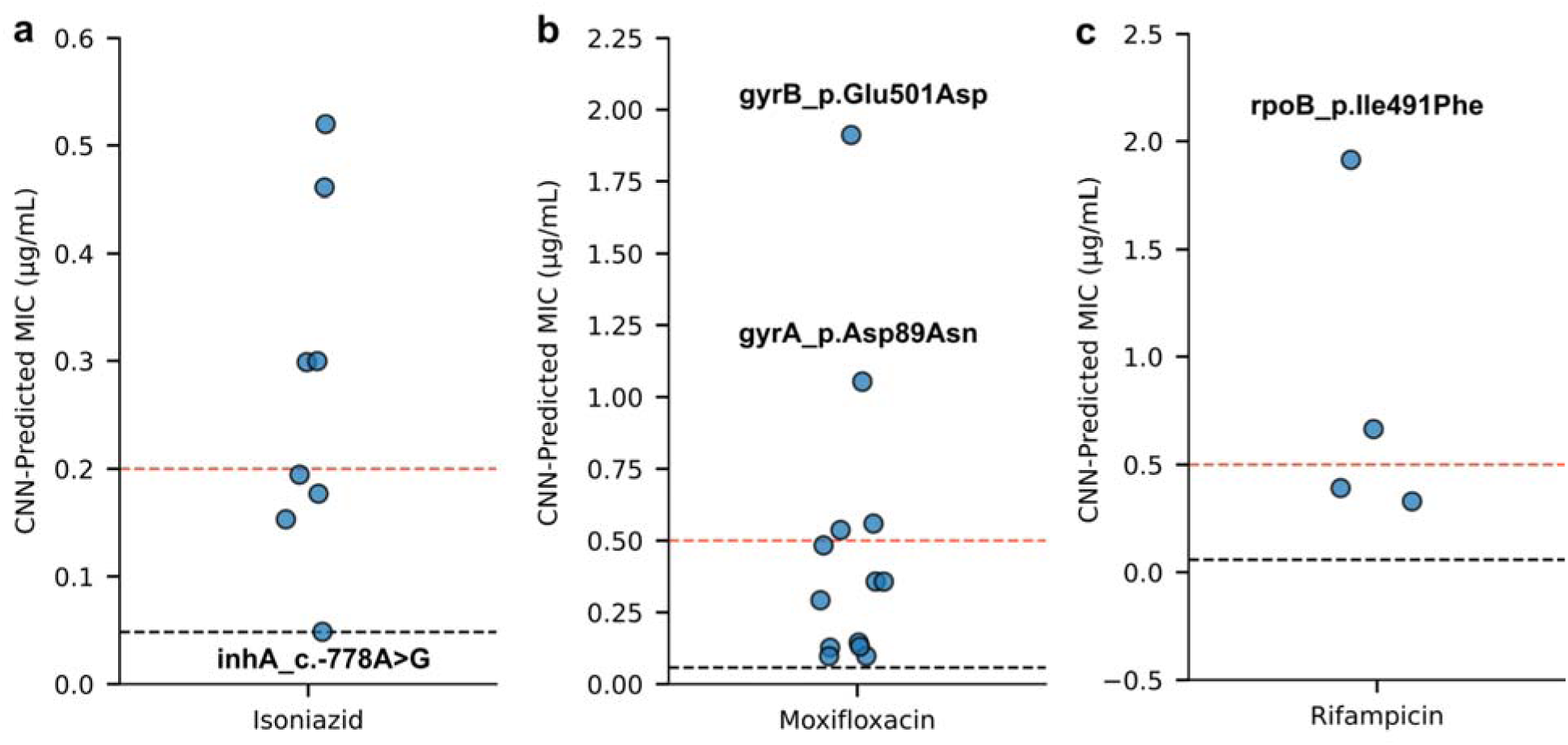
*In silico* MIC predictions for mutations classified as borderline or intermediate resistance in the WHO *Mtb* resistance mutation catalog. **a:** isoniazid, **b:** moxifloxacin, **c:** rifampicin, with certain labeled mutations. Black lines are H37Rv predictions, and red lines are the critical concentrations for each drug. Intermediate resistance is defined as the 0.2 - 1 µg/ml for isoniazid. For moxifloxacin and rifampicin, intermediate ranges are not well-defined, but assumed variably to range 0.25 - 2 µg/mL in research studies for both drugs.^83,84^ Source data are in **Supplementary Data 4.**

**Extended Data Figure 6.**
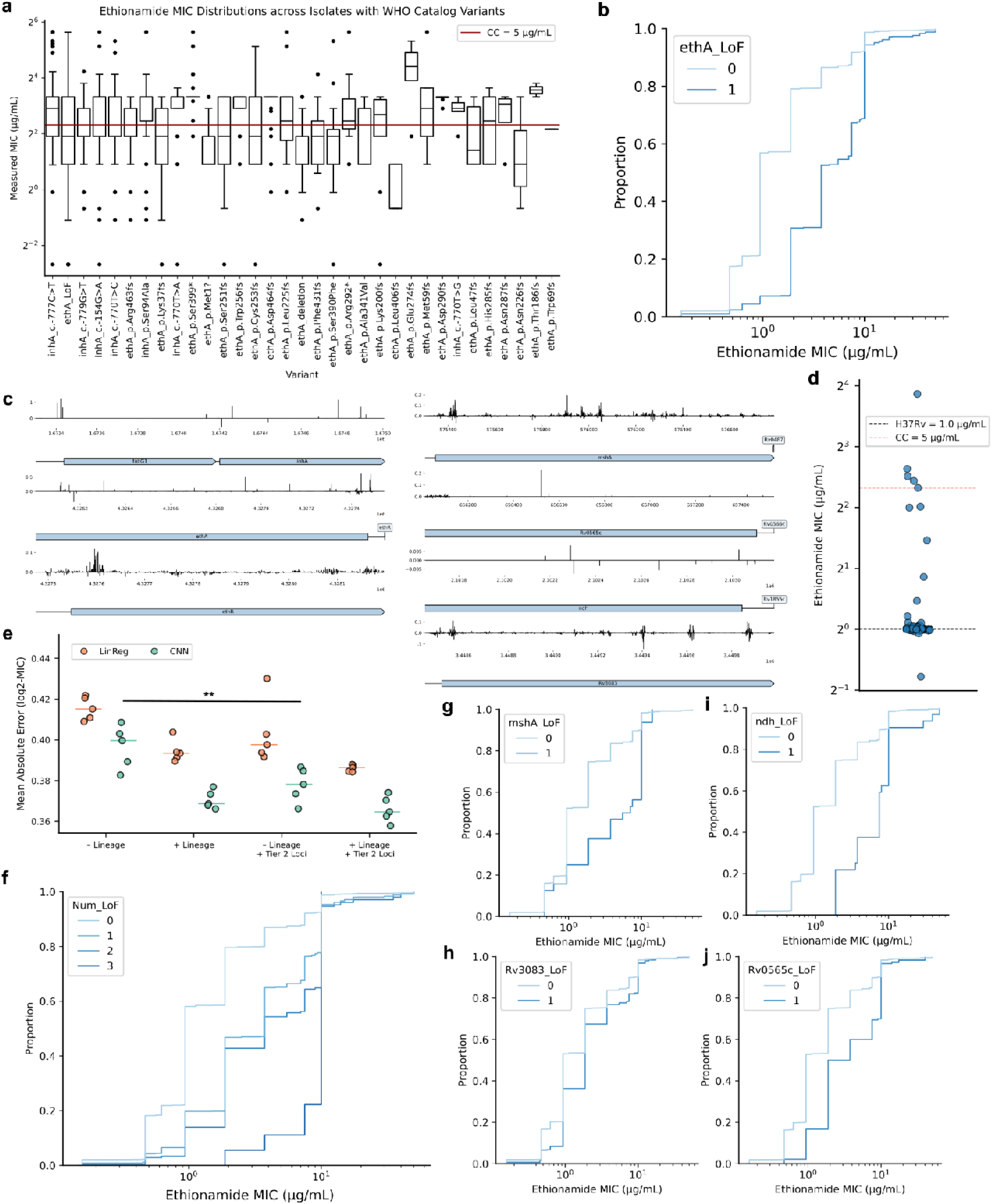
Mutations in non-ethAR genes are associated with ethionamide resistance. LoF is defined as frameshift variant, start lost, stop gained, or gene ablation. **a:** Measured MIC distribution of training set isolates containing ethionamide resistance-associated mutations from the WHO *Mtb* mutation catalog,^35^ showing a range of MICs across the susceptible and resistant ranges. **b:** Empirical cumulative distribution function (ECDF) of MIC between 10,120 isolates stratified by presence of any *ethA* LoF mutations. Critical concentration is 5 µg/ml. **c:** Saliency scores for all genes in the expanded ethionamide model. **d:** Strip plot of predicted MICs for 145 non-silent *mshA* mutations from the catalog, all of which are graded “Uncertain” in the catalog. **e:** Mean absolute errors of original and expanded ethionamide models, with and without the lineage barcode. Bar with asterisk indicates statistical significance at p<0.05 **f:** ECDF of MIC stratified by the number of genes among *ethA, mshA, ndh, Rv3083,* and *Rv0565* with LoF mutations. The average MIC of isolates with N genes with LoF mutations is significantly greater than the average MIC of isolates with N - 1 genes with LoF mutations, for N in [1, 2, 3] (one-sided Welch’s t-test, p ≤ 0.03). **g-j:** ECDFs of MIC between 10,120 isolates stratified by presence of any LoF mutations in *mshA* **(g),** *Rv3083* **(h),** *ndh* **(i),** or *Rv0565c* **(j)**.

**Extended Data Figure 7.**
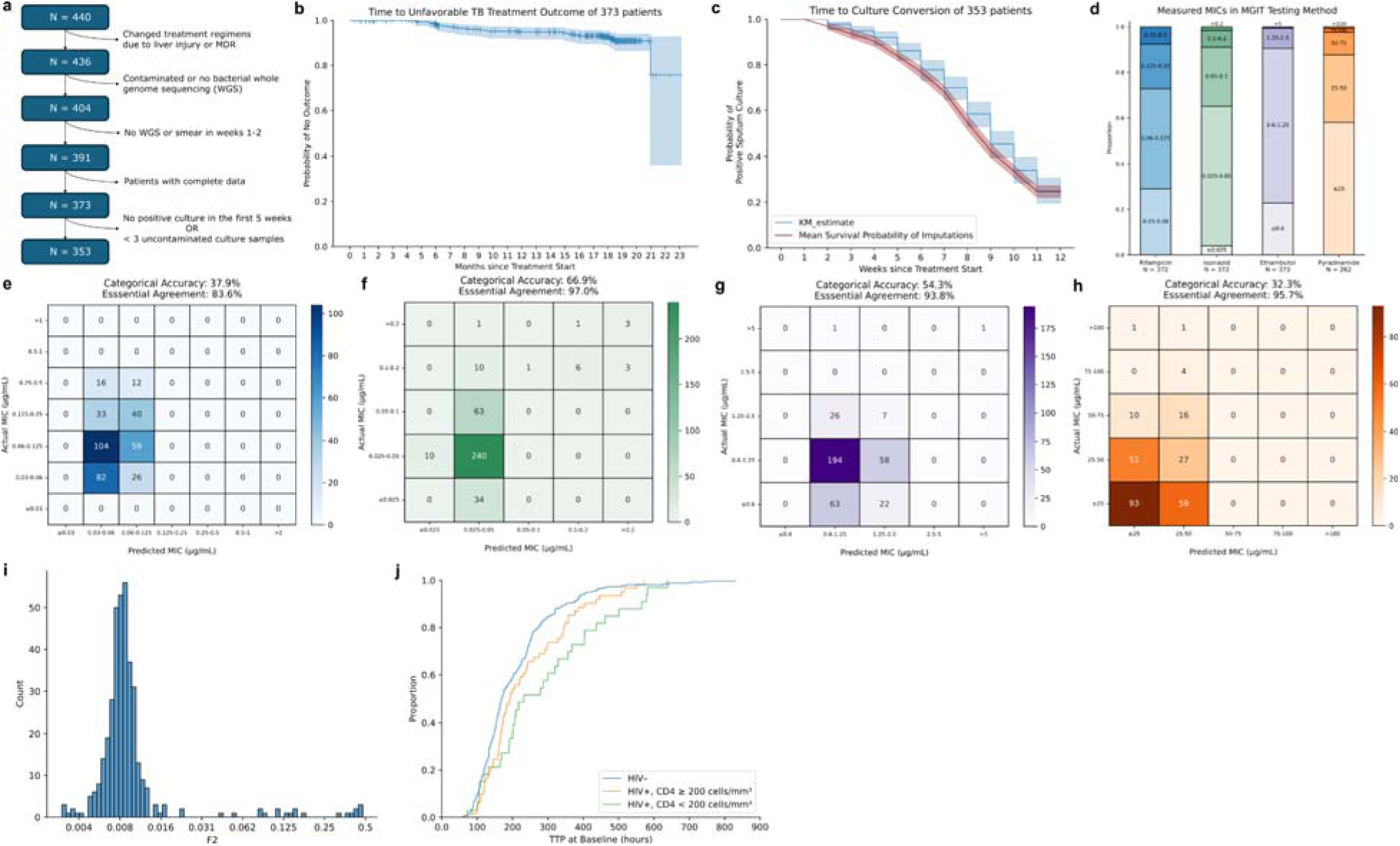
Participant characteristics and comparison of predicted and measured MICs in the TRUST cohort. **a:** Patient inclusion flowchart. **b:** Kaplan-Meier estimate of the survival function of unfavorable outcome-free survival up to 2 years after starting treatment. **c:** Kaplan-Meier estimates of the survival function of TCC across 12 weeks on the original data (blue) and the survival function averaged across 30 imputed culture datasets (red) with a 95% Wald confidence interval. **d:** Distribution of measured MICs in MGIT medium for the first-line drugs for 373 patients. **e-h:** Plots showing concordance between predicted and measured MICs for rifampicin **(e),** isoniazid **(f)**, ethambutol **(g)**, and pyrazinamide **(h)**. All measured and predicted MICs for the TRUST data are in MGIT medium. **i:** F2 score distribution for the baseline WGS samples for the 373 participants. **j:** Empirical cumulative distribution function (ECDF) of time to culture positivity (TTP) stratified by HIV and CD4 cell count status.

**Extended Data Figure 8.**
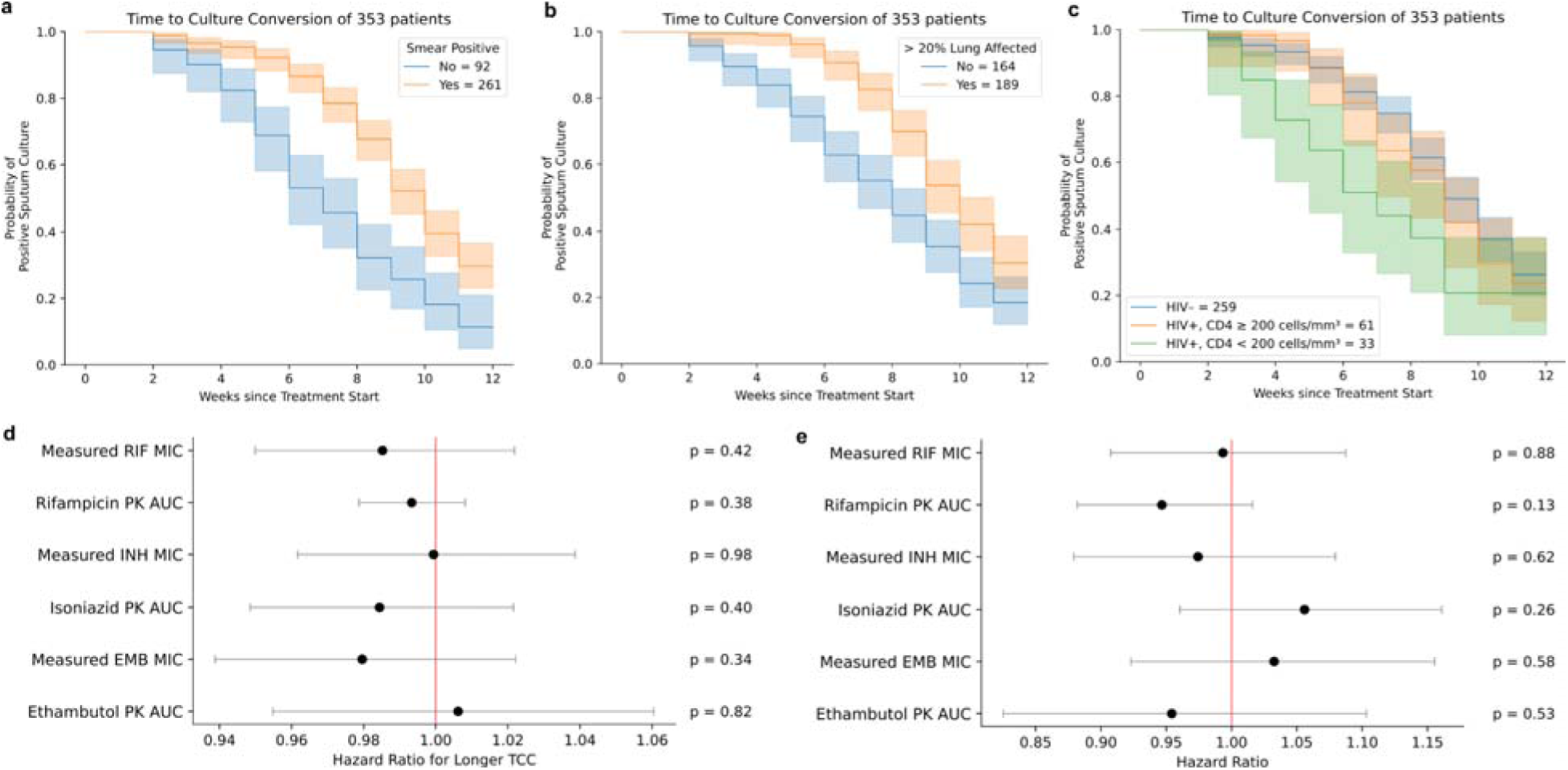
Additional Cox model and Kaplan-Meier estimate results. **a-c:** Kaplan-Meier estimates of TCC stratified by smear positivity **(a),** >20% lung involvement on chest X-ray **(b),** and HIV/CD4 count status **(c).** Forest plots of associations between measured MICs and TCC **(d)** or unfavorable outcomes **(e)** in multivariate models with measured MICs, predicted drug exposure, and patient characteristics. Hazard ratios for MIC variables are per increase in 0.1 µg/mL. PK AUC: Pharmacokinetic (PK) area under curve (AUC) of drug concentration over time. These values were predicted from patient demographic data.^76^

**Extended Data Figure 9.**
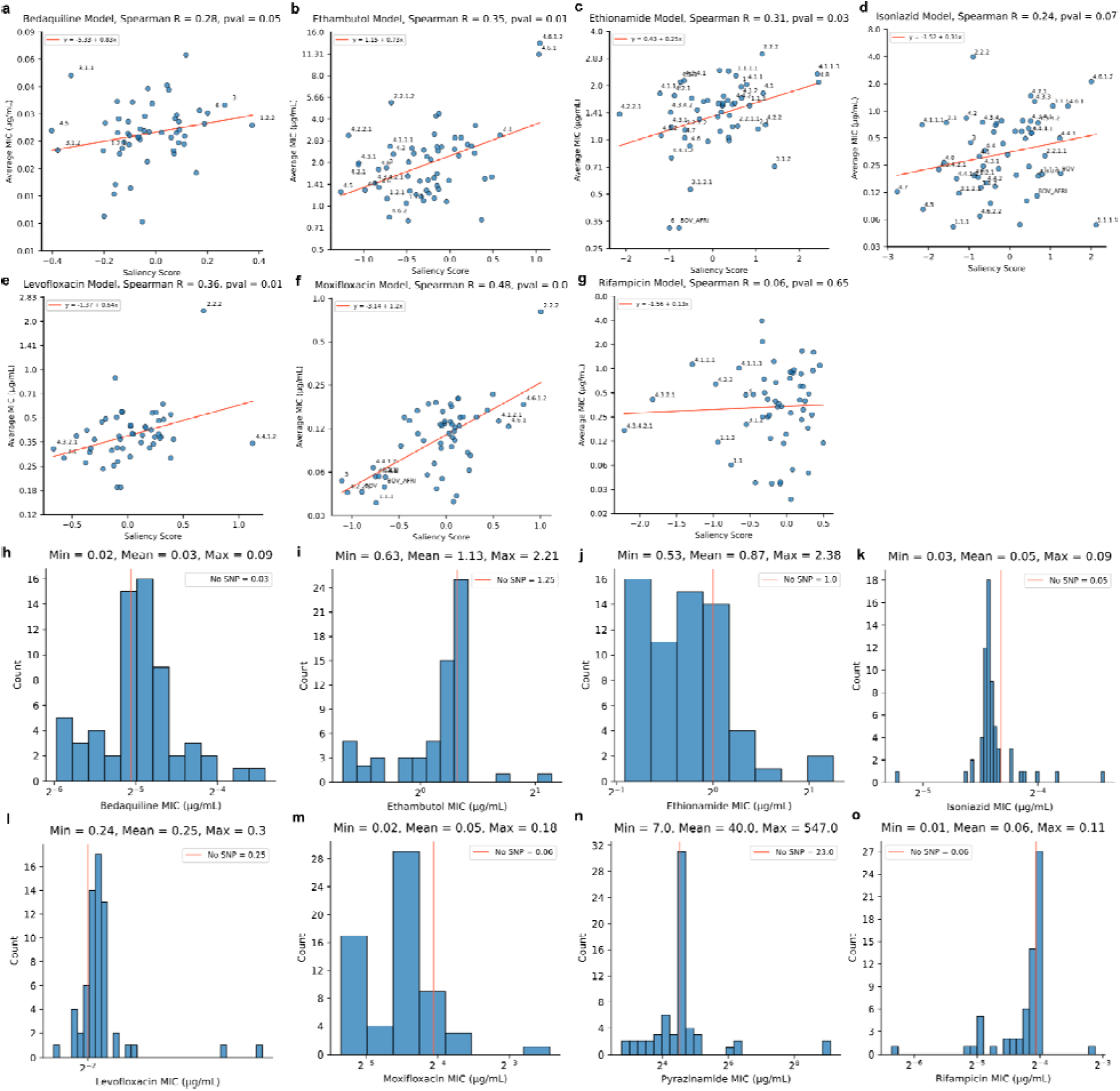
Additional lineage modeling results. **a-g:** Scatter plot of lineages and sublineages from the Coll 2014 scheme with at least 2 isolates and significant saliency scores for all single-drug models except pyrazinamide. The y-axis is the average MIC of training set isolates with each lineage SNP, and the x-axis is the saliency score. Lineages with a saliency score with an absolute value greater than 0.5 are annotated by name (greater than 0.2 for bedaquiline). Spearman correlations with p-values and linear regression lines of best fit are shown in the individual panels. **h-o:** Distributions of *in silico* predicted MICs for 62 lineage SNPs in for the 8 drug models with lineage SNPs. The minimum, mean, and maximum are given for each drug. The prediction for the H37Rv reference strain (no SNP present) is plotted as a red vertical line in each plot.

**Extended Data Figure 10.**
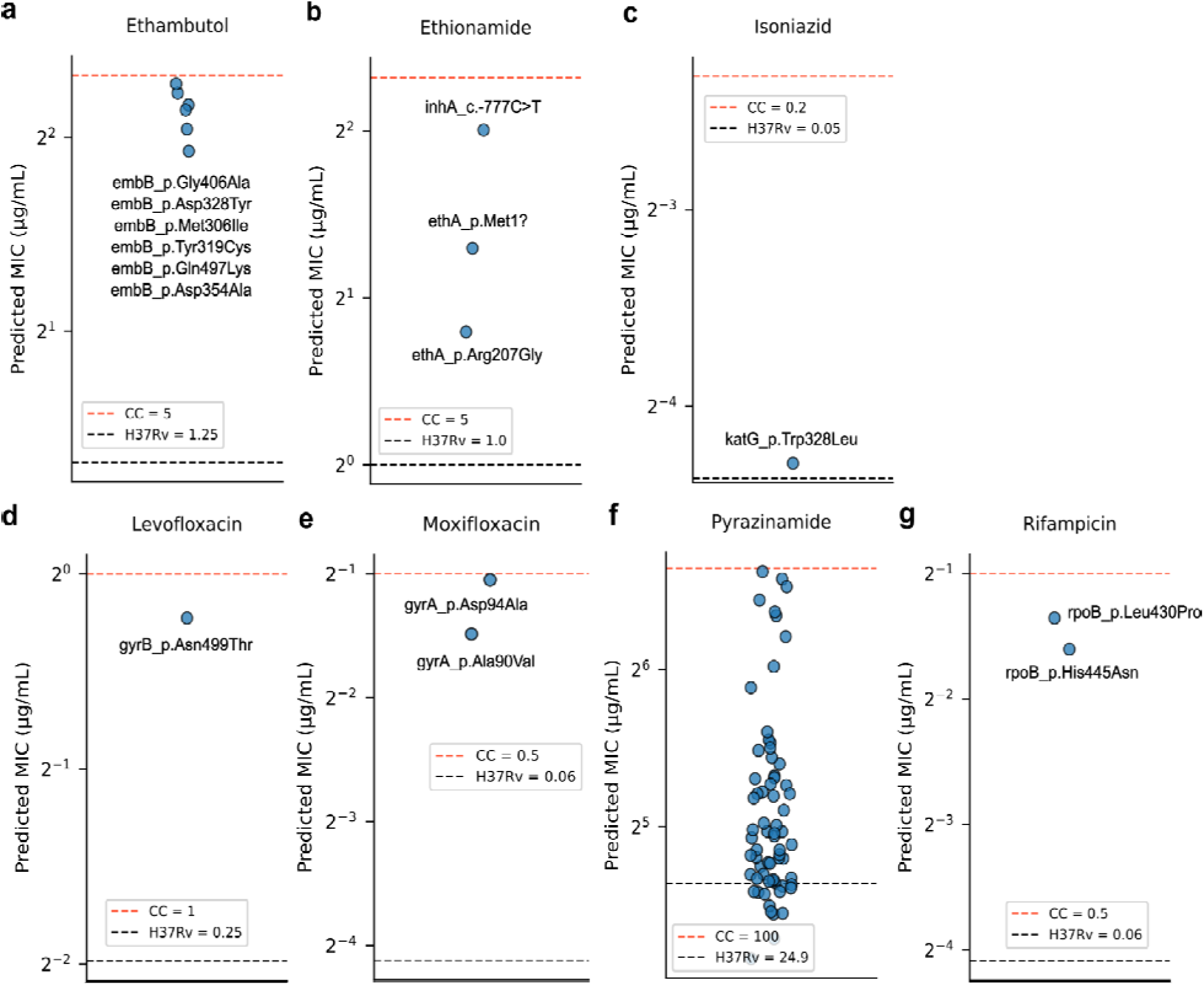
Group 1 resistance mutations from the WHO catalog that were not predicted to be associated with resistance by quantitative CNNs. Each panel is the MIC distribution of missed mutations for a single drug. **a-g:** in order, ethambutol, ethionamide, isoniazid, levofloxacin, moxifloxacin, pyrazinamide, and rifampicin. Certain mutations are annotated. Black dashed lines are the MIC predictions for H37Rv, and red dashed lines are the critical concentrations (CC) for each drug.

## Data

**<u>Supplementary Data 1:</u>** Isolate IDs, lineages and F2 strain mixing metrics, and MICs for 8 drugs, including the bedaquiline augmented and pyrazinamide binary datasets, which are separate sheets. All MICs are in 7H10 medium, except for bedaquiline (7H11) and pyrazinamide (MGIT), and both the MIC bounds and midpoint are listed. The train-validation-test splits are also given. Each sheet corresponds with a single drug dataset. Samples that have since been made publicly available have a public ID in the BIOSAMPLE_ACCESSION column.

**<u>Supplementary Data 2:</u>** List of data contributors, affiliations, and email addresses from the MIC-ML consortium, who provided genotypic and phenotypic data for model training.

**<u>Supplementary Data 3:</u>** Model performances for 5 cross-validation splits for both CNN and L2 regression for all drugs, with and without lineage SNPs, amino acid properties (without amino acid properties only for pyrazinamide), additional tier 2 loci (ethionamide only), and augmentation (bedaquiline only). Both binned mean error and mean error computed relative to MIC midpoints are listed for each quantitative model. The binary pyrazinamide model results are also in this table (Model = “Binary CNN” or “LogReg”) with AUCs.

**<u>Supplementary Data 4:</u>** *In silico* MIC predictions for mutations from the WHO catalog. Sheet 1: all predictions for quantitative models. Sheet 2: *In silico* predictions for mutations from the WHO catalog by the binary pyrazinamide model. Predictions are resistance probabilities, which have been binarized to a resistant vs. susceptible label. Sheet 3: mutations not associated with resistance based on literature curation for the 2^nd^ version of the WHO mutation catalog. Sheet 3: 28 mutations with literature evidence against association with resistance. Sheet 4: 82 Group 1 mutations with predicted MICs below the respective critical concentrations. Sheet 5: 25 mutations associated with low-level resistance to isoniazid, moxifloxacin, and rifampicin.

**<u>Supplementary Data 5:</u>** 247 nucleotide sites across all 8 drugs (244 unique sites due to locus redundancy across some drugs) newly associated with resistance (in the 70^th^ score percentile or higher within a single model, occur in at least 5 isolates, and are not in the WHO catalog.

**<u>Supplementary Data 6:</u>** Binary classification metrics (sensitivity, specificity, and F1 score) between the WHO catalog classification method and the quantitative CNNs (or binary, in the case of pyrazinamide). The “_LB” and “_UB” suffixes indicate lower and upper bounds of 95% exact binomial confidence intervals computed using the Clopper-Pearson method.

**<u>Supplementary Data 7:</u>** Site-saturation mutagenesis results for pncA, ethA, katG, gyrA, gyrB, and rpoB and Getis-Ord clustering scores (“G_score” column) for average predicted log_2_FC per site. For gyrA and gyrB, G scores for only residues solved in the crystal structure were computed. “BH_pval” = FDR-corrected p-value using Benjamini-Hochberg correction. “clustering_result” is 1 for hotspots, -1 for coldspot, and NA for all else. “average” is the average predicted log_2_FC for each codon.

**<u>Supplementary Data 8:</u>** All variants in genes graded in the 2023 WHO *Mtb* mutation catalog in the samples from participants in the TRUST study. Quality control fields output by the pilon variant calling software – FILTER, IMPRECISE, AF (allele fraction), DP (read depth), BQ (base quality), MQ (mapping quality), IC (reads supporting an insertion), and DC (reads supporting a deletion) – are included.

## Auxiliary

## Author Contributions Statement

S.G.K. implemented all code and ran all analyses, except for the Getis-Ord statistic computation, which was implemented by A.G.G. S.G.K. and M.R.F. interpreted the data and results and wrote the manuscript. B.C.M. and N.R. prepared whole-genome sequencing libraries and performed MIC testing. S.K.G., S. Malatesta, and N.C. cleaned patient data and shared results for the survival analyses. S. Mulaudzi cleaned and shared phenotypic data from the MIC-ML consortium. All authors provided feedback on the manuscript. M.R.F. supervised the research.

## Acknowledgements

We thank Marie Wijk and Paolo Denti for providing pharmacokinetic predictions for the patient outcomes analysis. We thank the study participants and study staff for making this work possible.

## Code and Data Availability

All code, original figures, and data files are available at https://github.com/sanju99/MtbQuantCNN.

